# Restoring STAR*D: A Reanalysis of Drug-Switch Therapy After Failed SSRI Treatment Using Patient-Level Data with Fidelity to the Original STAR*D Research Protocol

**DOI:** 10.1101/2025.02.10.25321991

**Authors:** Colin Xu, Thomas T. Kim, Irving Kirsch, Martin Plöderl, Jay D. Amsterdam, H. Edmund Pigott

**Author notes:** Corresponding author: Ed Pigott, PhD.

## Abstract

**Background:** The *Sequenced Treatment Alternatives to Relieve Depression* (STAR*D) trial was designed to give guidance in selecting the best next-step treatment for depressed patients who did not remit during their first, and/or subsequent, antidepressant trial, with up to four trials per patient. Our prior research documented protocol violations which inflated STAR*D’s reported cumulative remission rate by 91.4%. A similar reanalysis of the step-2 drug-switch trial has not been done until now.

**Methods:** We reanalyzed the patient-level dataset of STAR*D’s drug-switch treatment therapies—*with fidelity to the original research protocol and related publications*—to determine whether there were clinically-relevant differences in results compared to the original publication.

**Results:** While our reanalysis largely comported with STAR*D’s published findings of no significant differences between drug-switch treatments, we found the following discrepancies: Lower than reported step-2 remission rates ranging from 16.2 to 19.3% (versus 17.6 to 24.8%); A significant increase in treatment-emergent suicidal ideation during the step-2 drug-switch therapies ranging from 11.2 to 15.0% compared to step-1 citalopram treatment (9.0%); A four times greater number of severe suicidal behaviors reported by the treating clinicians compared to the published suicide-related Serious Adverse Events (16 versus 4); and A sustained remission rate of only 3.1 to 8.4%.

**Conclusion:** Compared to the original publication, our reanalysis found lower remission rates and more suicidal risk than reported. This adds to the discrepancies found in our prior reanalysis and also to the finding that switching antidepressants is not well supported by the evidence.

## Introduction

Funded by the National Institute of Mental Health (NIMH), the $35-million *Sequenced Treatment Alternatives to Relieve Depression* (STAR*D) trial is the largest and most influential, prospective comparative treatment effectiveness study ever conducted of outpatients with major depressive disorder (MDD).^1-7^

STAR*D provided up to four antidepressant treatment trials per patient and was designed to give guidance in selecting the best next-step treatment for the many patients who fail to gain sufficient relief from their first, and/or subsequent, antidepressant trial(s).^8^ To mimic clinical practice, STAR*D used an open-label research design, enrolled patients seeking routine medical or psychiatric care, and included patients with a wide range of common co-morbid medical and psychiatric conditions to enhance the generalizability of findings to real-world practice.

In step-1, 4041 patients who screened positive for MDD were prescribed citalopram as their first treatment.^1^ The STAR*D investigators developed a system of measurement-based care to guide increasingly aggressive antidepressant medication dosing to “*ensure that the likelihood of achieving remission was maximized and that those who did not reach remission were truly resistant to the medication*.”^1, p.30^ Patients who failed to gain adequate relief from citalopram could select acceptable treatment options for randomization in step-2.

In step-2, patients had seven treatment options to choose from: four switch options in which citalopram was stopped and the new treatment started and three augmentation options in which citalopram was combined with a second antidepressant treatment. Cognitive therapy was available as either a switch or augmentation option.

Exceedingly few patients accepted randomized assignment for all treatment options in step-2, with the vast majority opting to either have their medication switched or augmented.^2,3^ The investigators reported that patients who opted to switch their medication were more severely ill entering step-2, and had higher rates of citalopram intolerance in step-1, than patients who opted to augment their citalopram.^2^

Given these differences, the STAR*D investigators analyzed drug-switch and drug-augmentation groups separately in the published articles.^2,3^

The three switch medications were pharmacologically-distinct from one another. They were: (1) sustained-release bupropion; (2) the SSRI sertraline; and (3) extended-release venlafaxine.

Based on STAR*D’s step-2 results, the investigators reported that these pharmacologically-distinct treatments “*did not differ significantly with respect to outcomes, tolerability, or adverse events*,”^2, p.1231^ and therefore provided no evidence-based guidance for selecting the best next-step drug-switch treatment option after a failed response to initial citalopram therapy.

A reanalysis of STAR*D’s step-2 drug-switch dataset is warranted for three reasons

First, our previous reanalysis under the *Restoring Invisible and Abandoned Trials* (RIAT) initiative discovered protocol violations which led to an overestimation of remission rates.^9^ Similar discrepancies could be present in STAR*D’s step-2 drug-switch treatments and should be explored.

Second, long-term effectiveness (up to one year) results in step-2 switch therapies remain unpublished despite STAR*D’s primary investigator (PI) stating in 2007 that STAR*D investigators would “*compare longer-term outcomes of the various treatments*.”^10, p.201^

Third, the extent of treatment-emergent suicidal ideation (TESI) during step-2 drug-switch treatments remains unpublished. Starting and stopping antidepressant medication has been shown to decrease the likelihood of remission to the next treatment trial,^11-14^ but increases the risk of suicide.^15^ Furthermore, an analysis of the Food and Drug Administration Adverse Event Reporting System found that drugs associated with TESI are also linked to increased suicide attempts and suicides.^16^

We believe that these data will better inform clinicians of the sustainability of treatment gains and risks; and are therefore vital for them in determining the best next-step treatment option after a patient’s failed response to SSRI therapy.

The current RIAT study reanalyzed STAR*D’s step-2 drug-switch trial with fidelity to the original STAR*D research protocol and related publications using the patient-level dataset downloaded from NIMH in January 2025. In addition to comparing outcomes using STAR*D’s primary measure (i.e., the Hamilton Rating Scale for Depression (HRSD)), we also compared: (a) outcomes for patients who entered the one-year follow-up phase; (b) incidents of severe suicidal ideation/suicide attempts during step-2 drug-switch treatment; and (c) TESI rates using the same methodology STAR*D investigators used in step-1.^17^

## Methods

### Patients

STAR*D patients were 18 to 75 years old and seeking care at 18 primary and 23 psychiatric clinics. Details are provided elsewhere.^1^

In step-2, 580 drug-switch patients met the inclusion criteria for data analysis. Supplement 1 presents the number of drug-switch patients who were excluded in our reanalysis, yet included in the

STAR*D investigators’ step-2 article, and the reasons for their exclusion. Supplement 2 is the patient flow chart moving from step-1 citalopram treatment to the step-2 drug-switch therapies.

### STAR*D treatment and assessment of outcomes: original analysis and our RIAT reanalysis

STAR*D investigators sought to provide the highest quality of acute and continuing-care treatment to maximize the number of remissions while minimizing relapse and dropouts (see Supplement 3). Each of the four treatment steps consisted of 12 weeks of antidepressant therapy, with an additional 2 weeks for patients deemed close to remission. All patients received citalopram as their step-1 treatment.

Treatment was administered using a system of measurement-based care that assessed symptoms using the Quick Inventory of Depressive Symptoms - Self-Report and Clinician versions (QIDS-SR and QIDS-C), side effects, and medication adherence at each clinic visit. Patients without a satisfactory clinical outcome in step-1 had the option to enter step-2 and either ‘switch’ or ‘augment’ their citalopram treatment. Patients who switched treatments stopped citalopram without a washout period before starting the new medication.

In the current analysis, we focused on patients who switched from citalopram in step-1 to another antidepressant in step-2 (see Supplement 4 for a description of the medications and their dosing).

STAR*D investigators implemented an “equipoise-stratified randomized” research design in which patients could opt to not receive certain step-2 treatment options.^18^ While patients were strongly encouraged to accept all seven step-2 treatments for randomization, only 4.0% accepted randomization to all drug-switch and augmentation options, preventing a head-to-head comparison between drug-switch/drug-augmentation treatments.^2^

### Primary Outcome – Symptom Remission

The primary outcome according to the STAR*D’s protocol was symptom remission, defined by a score of <8 on patients’ step-2 exit HRSD. The HRSD was obtained at treatment entry and exit by research outcome assessors (ROAs) blind to treatment status. STAR*D investigators prespecified that “*primary analyses classified patients with missing exit HRSD scores as nonremitters a priori*.”^1, p.34^ In our RIAT reanalysis, we analyze the primary outcome data using these prespecified criteria.

Due to the high rate of drug-switch patients missing an exit HRSD (n = 170; see Supplement 5), we conducted a sensitivity analysis where we imputed HRSD scores though mapping their final QIDS-SR score to the HRSD to compute secondary outcome measures for remission, response (defined as a ≥ 50% reduction in HRSD), and mean symptom improvement. This is the same imputation method used by the STAR*D investigators.^2^

### Suicide outcomes: TESI and Serious Adverse Events (SAE) involving suicidality

TESI was defined by the STAR*D investigators as a score of ≥2 on any postbaseline visit for patients whose baseline score was <2 on the QIDS-C suicide item.^17^ The QIDS-C suicide item is scored: “(0) Does not think of suicide or death; (1) Feels life is empty or is not worth living; (2) Thinks of suicide/death several times a week for several minutes; and (3) Thinks of suicide/death several times a day in depth, or has made specific plans, or attempted suicide.”^19^ For our RIAT-analysis, we use these same criteria to analyze TESI.

The original publication also reported on suicide-related SAEs. For our RIAT-analysis, we examined the patient-level SAE data files for those patients whose treating clinician rated them as 3, “*Thinks of suicide/death several times a day in depth, or has made specific plans, or attempted suicide*” on the QIDS-C scale. These clinician-rated incidents of severe suicidality were then compared to suicide-related SAEs reported by the STAR*D investigators.^2^

### One year follow-up treatment for responders and remitters

According to the STAR*D study plan, patients scoring <6 on their last clinic-administered QIDS-C during step-2 treatment were encouraged to enter the 12-month follow-up phase. A QIDS score <6 was estimated by STAR*D investigators to correspond to a HRSD score of <8, STAR*D’s definition of remission.^20^ Clinicians strongly encouraged patients who did not obtain a QIDS-C-defined remission to enter the next-step treatment. However, patients who failed to attain a QIDS-C-defined remission, but had a ≥50% reduction on the QIDS-C and refused randomization to a next-step treatment, were encouraged to enter follow-up.

During the follow-up phase, patients continued their “*previously effective acute treatment medication(s) at the doses used in acute treatment but that any psychotherapy, medication, or medication dose change could be used*.”^7, p.1908^

During follow-up, the STAR*D ROAs collected HRSD data from patients at months 3, 6, 9, and 12. A telephonic integrated voice response version of the QIDS (QIDS-IVR) was also collected at months 1, 2, 4, 5, 7, 8, 10, and 11. Missing quarterly HRSD scores were imputed by mapping the closest available QIDS-IVR score ± 4 weeks to that visit timepoint.

Relapse was defined by the STAR*D investigators as an HRSD score ≥14 at one of the four follow-up timepoints (months 3, 6, 9, and 12). For patients who met remission criteria and entered follow-up, we calculated the number of patients who sustained their remission (i.e., HRSD score <8 at all four timepoints). We also calculated a lenient definition of sustained remission. Patients met this lenient definition if they had HRSD scores <8 at one or more timepoints, but had missing data at the other timepoints.

### Statistical methods

Drug-switch treatments were compared with χ^2^-tests for discrete outcomes (remission, response, TESI, relapse and sustained remission) and ANOVA for continuous outcomes. Step-2 remission rates as calculated in our RIAT reanalysis were compared against those reported in the original STAR*D publication using χ^2^ tests.^2^

Supplement 6 provides the statistical code that the first and second authors used to analyze the STAR*D patient-level dataset from the NIMH-provided datasets.

## Results

### Patient characteristics

Demographic and clinical characteristics of the 580 patients who switched drugs in step-2 and met inclusion criteria for data analysis are described in Supplement 7.

### Step-2 drug-switch outcomes

Table 1 presents the acute care outcomes.

**Table 1:**
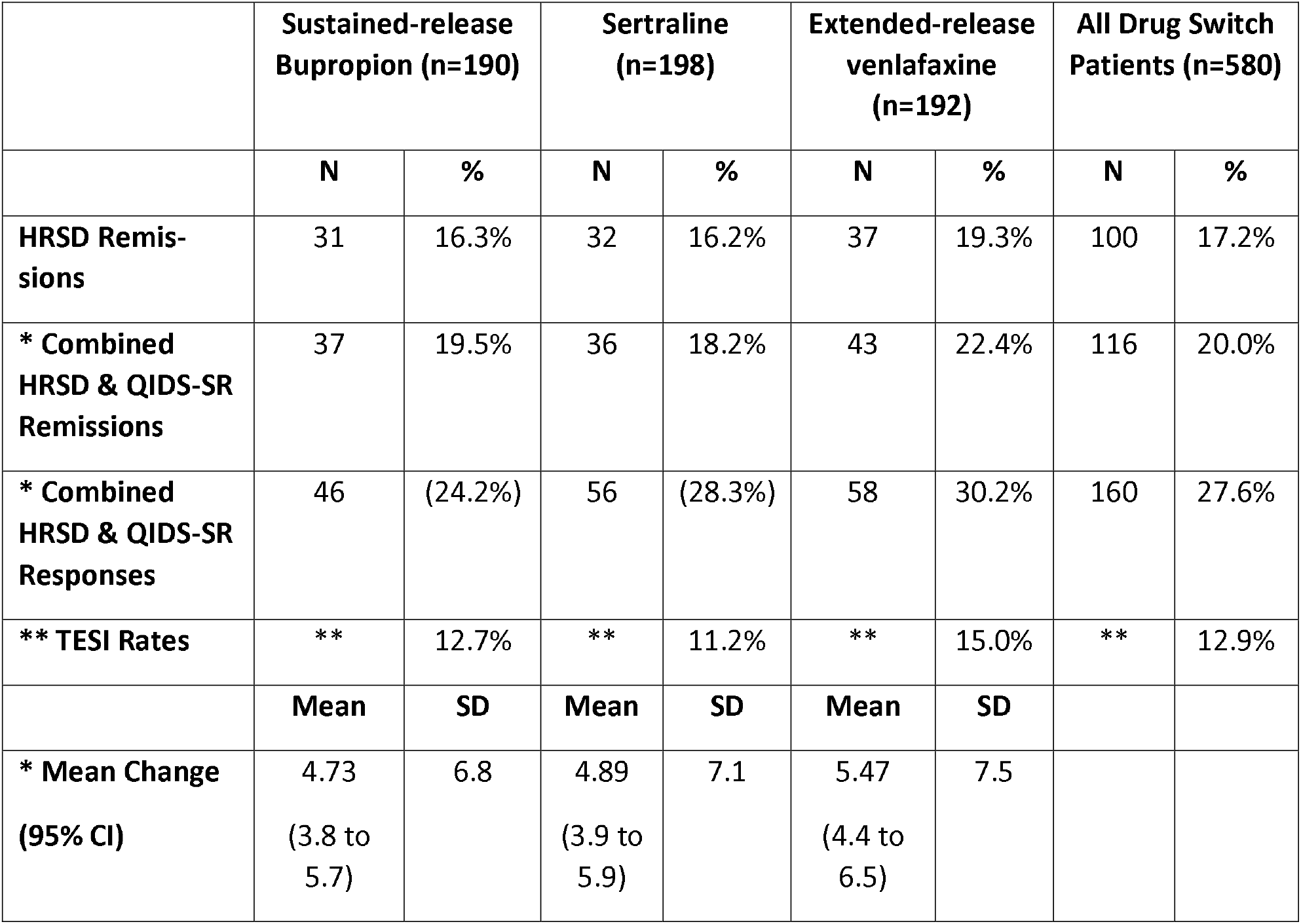
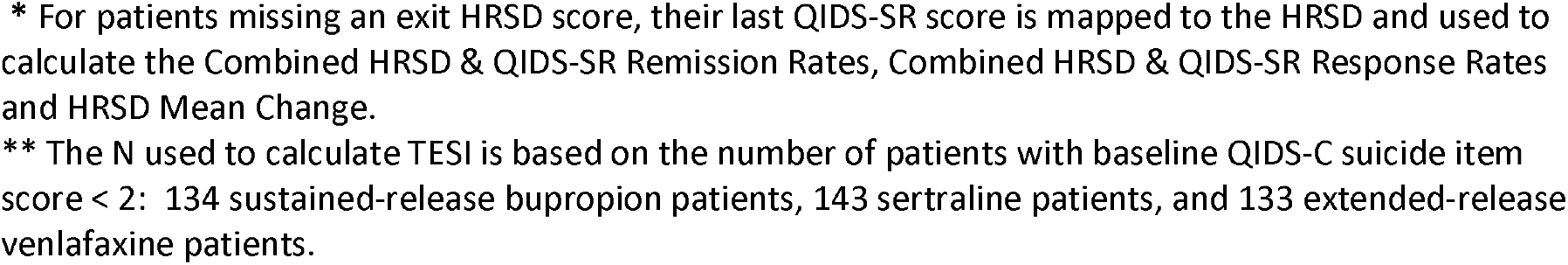
Acute Care Outcomes for Drug-Switch Patients.

|  | **Sustained-release**<br>**Bupropion (n=190)** |  | **Sertraline**<br>**(n=198)** |  | **Extended-release**<br>**venlafaxine**<br>**(n=192)** |  | **All Drug Switch**<br>**Patients (n=580)** |  |
| --- | --- | --- | --- | --- | --- | --- | --- | --- |
|  | **N** | **%** | **N** | **%** | **N** | **%** | **N** | **%** |
| **HRSD Remissions** | 31 | 16.3% | 32 | 16.2% | 37 | 19.3% | 100 | 17.2% |
| **\* Combined**<br>**HRSD & QIDS-SR**<br>**Remissions** | 37 | 19.5% | 36 | 18.2% | 43 | 22.4% | 116 | 20.0% |
| **\* Combined**<br>**HRSD & QIDS-SR**<br>**Responses** | 46 | (24.2%) | 56 | (28.3%) | 58 | 30.2% | 160 | 27.6% |
| **\*\* TESI Rates** | ** | 12.7% | ** | 11.2% | ** | 15.0% | ** | 12.9% |
|  | **Mean** | **SD** | **Mean** | **SD** | **Mean** | **SD** |  |  |
| **\* Mean Change**<br>**(95% CI)** | 4.73<br>(3.8 to<br>5.7) | 6.8 | 4.89<br>(3.9 to<br>5.9) | 7.1 | 5.47<br>(4.4 to<br>6.5) | 7.5 |  |  |

\* For patients missing an exit HRSD score, their last QIDS-SR score is mapped to the HRSD and used to calculate the Combined HRSD & QIDS-SR Remission Rates, Combined HRSD & QIDS-SR Response Rates and HRSD Mean Change.
\*\* The N used to calculate TESI is based on the number of patients with baseline QIDS-C suicide item score < 2: 134 sustained-release bupropion patients, 143 sertraline patients, and 133 extended-release venlafaxine patients.

There was no significant difference in the proportions of patients who met remission criteria between the three drug-switch treatments (χ^2^ [2, n = 580] = 0.83, *p* = 0.66). 16.3% of the 190 patients who received sustained-release bupropion remitted; 16.2% of the 198 patients who received sertraline remitted; and 19.3% of the 192 patients who received extended-release venlafaxine remitted.

Similarly, we found no significant differences on the secondary outcome measures for remission (χ^2^ [2, *n* = 580] = 1.1.3, *p* = 0.568); response (χ^2^ [2, *n* = 580] = 1.79, *p* = 0.41), nor a statistically significant difference in mean symptom improvement (p = 0.57).

We found substantially lower remission rates in our RIAT analysis compared to those reported by the STAR*D investigators (Fig. 1, Supplement 8). The difference in proportions of remission on the HDRS (primary outcome) in the original publication versus our reanalysis was just below statistical significance (21.2% vs 17.5%; χ^2^ [1, *n* = 1307] = 3.42, *p* = 0.06).

**Figure 1:**
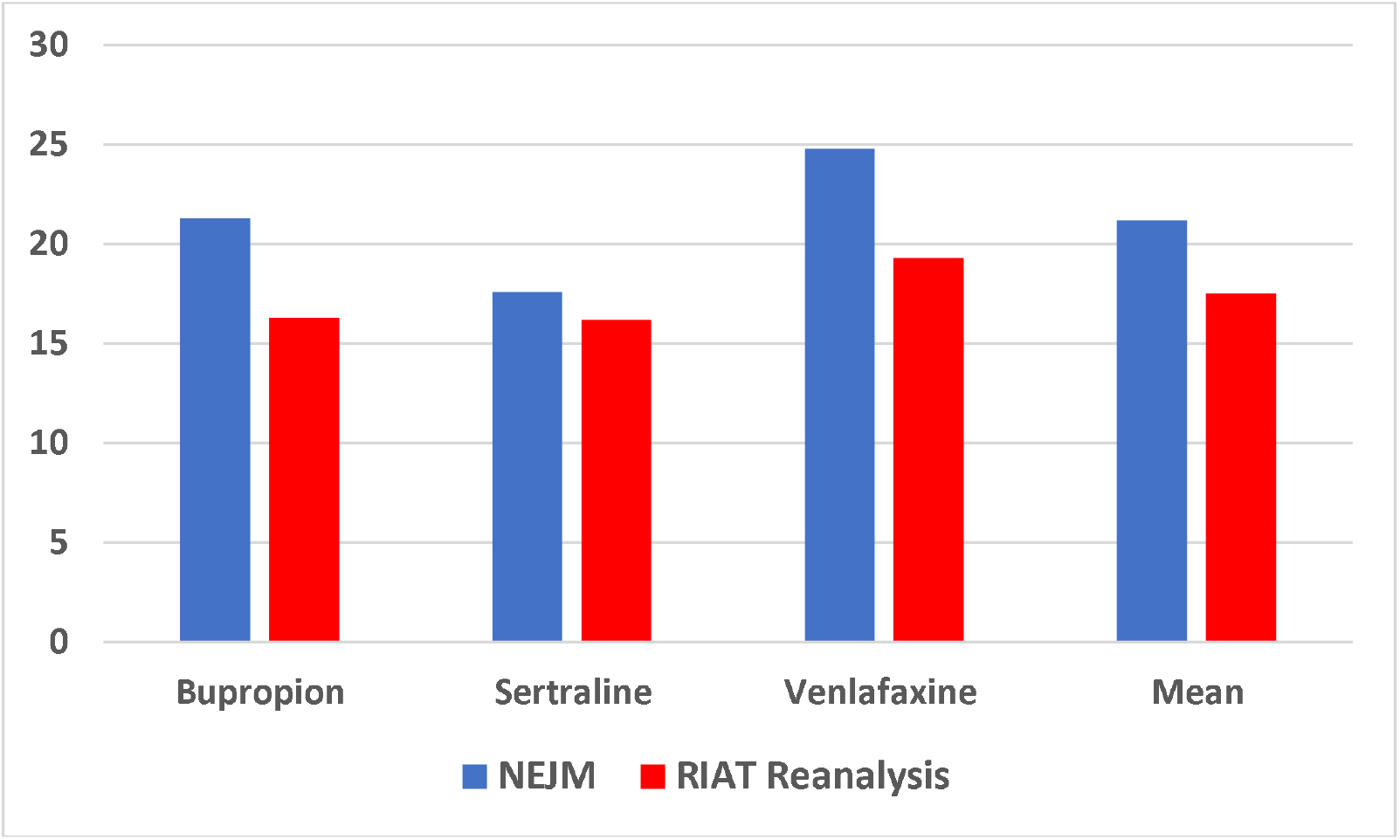
Reported Drug-Switch HRSD Remission Rates.

### TESI rates for step-2 drug-switch patients and SAEs involving severe-suicidality

The TESI rates for drug-switch patients were 12.7% for sustained-release bupropion, 11.2% for sertraline, and 15.0% for extended-release venlafaxine and did not differ significantly across treatment groups (χ^2^ [2, *n* = 410] = 0.92, *p* = 0.63).

There was a significantly greater proportion of patients with TESI in step-2 drug-switch treatment than step-1 treatment (12.9% vs 9.0%; χ^2^ [1, *n* =2533] = 6.27, *p* < 0.01, Supplement 9) and averaged a 30% increase in TESI rates between step-1 and step-2 drug-switch treatments (see Figure 2).

**Figure 2:**
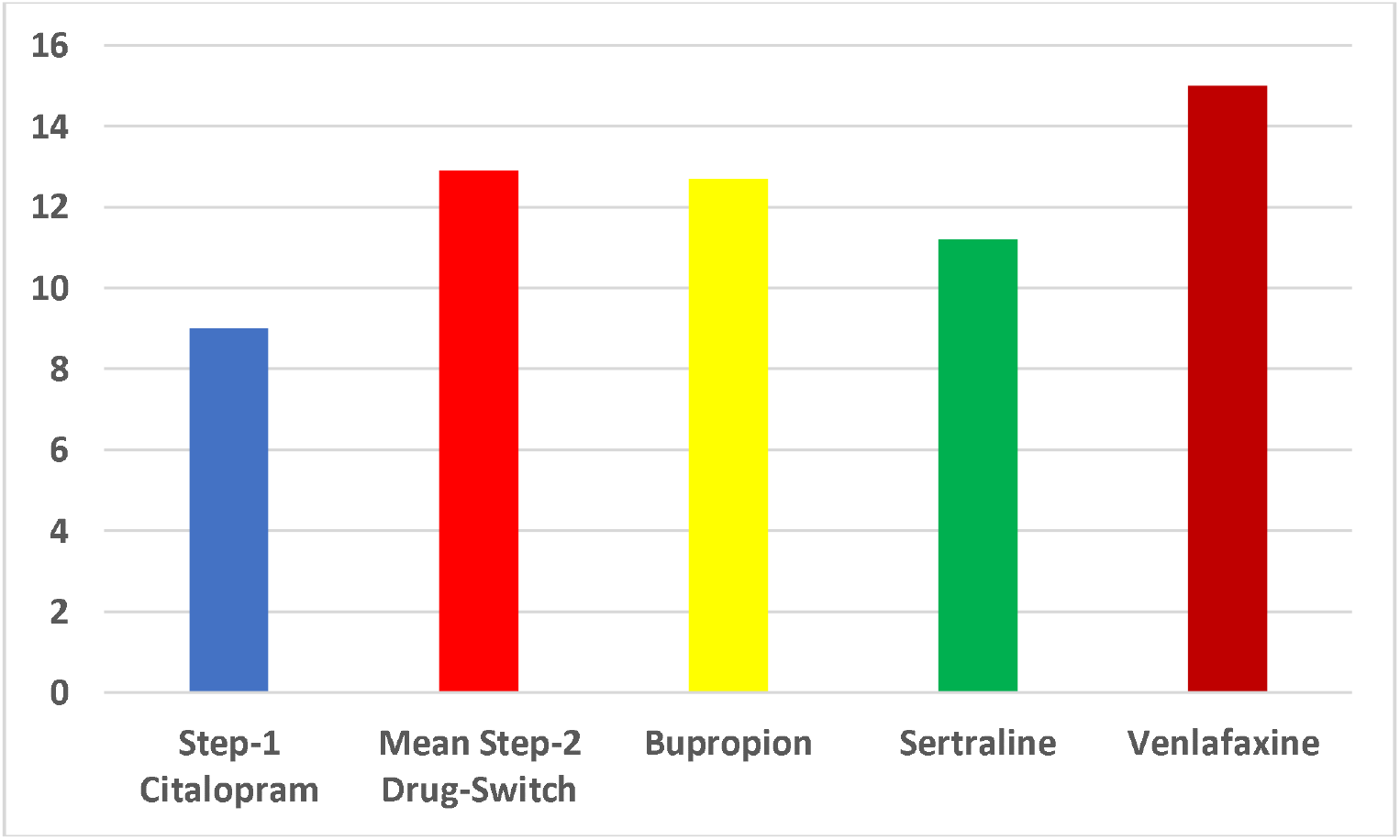
Rate of Treatment-Emergent Suicidal Ideation.

The STAR*D investigators reported four SAEs in which drug-switch patients had “Psychiatric hospitalization for suicidal ideation or attempt”, and no SAEs of patients who had “Suicidal ideation without hospitalization”—*a severe-suicidality rate of 0.55%*. In contrast, our RIAT reanalysis found that there were 16 instances in which clinicians rated their patients as “*Thinks of suicide/death several times a day in depth, or has made specific plans, or attempted suicide” on the QIDS-C suicide item*—a severe-suicidality rate of 2.8% as reported by treating clinicians. Only four of these patients had suicide-related SAEs recorded in the patient-level dataset. Table 2 compares suicide-related SAEs as reported in the original drug-switch article verses what was reported by the treating clinicians.

**Table 2:** Severe Suicidal Ideation and Attempts in Drug-Switch Patients as Reported in the Original Publication and Clinician-Rated QIDS Data in Our RIAT Reanalysis with 147 Fewer Patients.

|  | BUP<br>(N = 239) | SER<br>(N = 238) | VEN<br>(N = 250) | Total |
| --- | --- | --- | --- | --- |
| “Psychiatric hospitalization for suicidal ideation or attempt” as reported in NEJM based on 727 patients | 0 | 2 | 2 | 4 |
| “Suicidal ideation without hospitalization” as reported in NEJM based on 727 patients | 0 | 0 | 0 | 0 |
| # of times patients were rated by their treating clinicians as “*Thinks of suicide/death several times a day in depth, or has made specific plans, or attempted suicide*” on the QIDS-C suicide item in our RIAT reanalysis of the 580 drug-switch patients who meet STAR*D’s pre-specified inclusion for data analysis criteria | 5 | 6 | 5 | 16 |

### Follow-up phase: Relapse and sustained remission

Table 3 presents the relapse and sustained remission rates during follow-up.

**Table 3:**
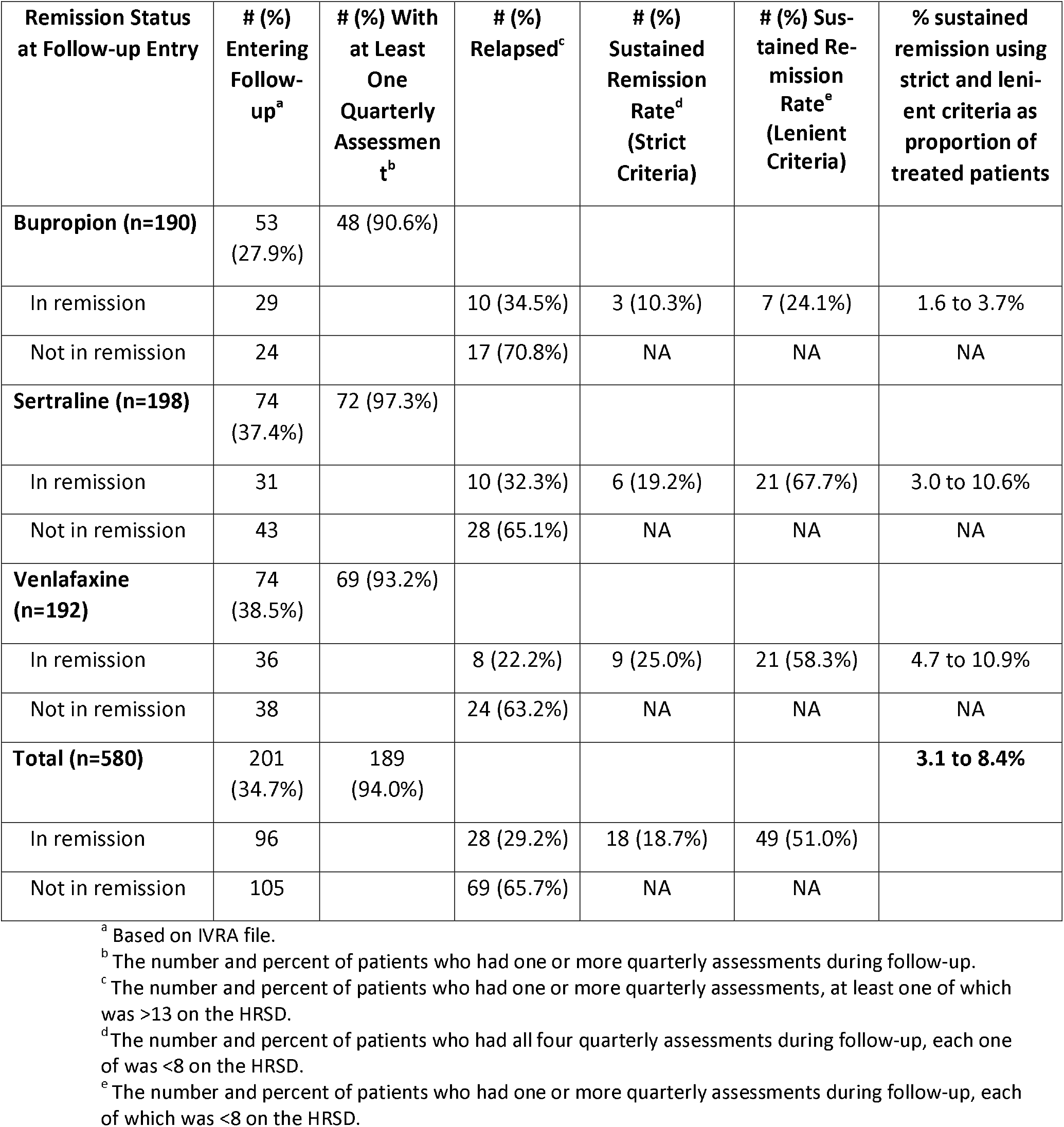
Relapse and Sustained Remission Rates During Follow-up.

| **Remission Status at Follow-up Entry** | **# (%) Entering Follow-up<sup>a</sup>** | **# (%) With at Least One Quarterly Assessment<sup>b</sup>** | **# (%) Relapsed<sup>c</sup>** | **# (%) Sustained Remission Rate<sup>d</sup> (Strict Criteria)** | **# (%) Sustained Remission Rate<sup>e</sup> (Lenient Criteria)** | **% sustained remission using strict and lenient criteria as proportion of treated patients** |
| --- | --- | --- | --- | --- | --- | --- |
| **Bupropion (n=190)** | 53 (27.9%) | 48 (90.6%) |  |  |  |  |
| In remission | 29 |  | 10 (34.5%) | 3 (10.3%) | 7 (24.1%) | 1.6 to 3.7% |
| Not in remission | 24 |  | 17 (70.8%) | NA | NA | NA |
| **Sertraline (n=198)** | 74 (37.4%) | 72 (97.3%) |  |  |  |  |
| In remission | 31 |  | 10 (32.3%) | 6 (19.2%) | 21 (67.7%) | 3.0 to 10.6% |
| Not in remission | 43 |  | 28 (65.1%) | NA | NA | NA |
| **Venlafaxine (n=192)** | 74 (38.5%) | 69 (93.2%) |  |  |  |  |
| In remission | 36 |  | 8 (22.2%) | 9 (25.0%) | 21 (58.3%) | 4.7 to 10.9% |
| Not in remission | 38 |  | 24 (63.2%) | NA | NA | NA |
| **Total (n=580)** | 201 (34.7%) | 189 (94.0%) |  |  |  | **3.1 to 8.4%** |
| In remission | 96 |  | 28 (29.2%) | 18 (18.7%) | 49 (51.0%) |  |
| Not in remission | 105 |  | 69 (65.7%) | NA | NA |  |

<sup>a</sup> Based on IVRA file.
<sup>b</sup> The number and percent of patients who had one or more quarterly assessments during follow-up.
<sup>c</sup> The number and percent of patients who had one or more quarterly assessments, at least one of which was >13 on the HRSD.
<sup>d</sup> The number and percent of patients who had all four quarterly assessments during follow-up, each one of which was <8 on the HRSD.
<sup>e</sup> The number and percent of patients who had one or more quarterly assessments during follow-up, each of which was <8 on the HRSD.

Overall, 29.2% of patients in the drug-switch treatments met remission criteria at entry into follow-up and relapsed (n=28). There was no significant difference between the drug-switch treatments (χ^2^ [2, *n* = 96] = 1.38, *p* = 0.50).

There was no significant difference in the proportions of patients who achieved a sustained remission between the three switch treatments when using the stringent criteria (χ^2^ [2, *n* = 96] = 2.28, *p* = 0.32). There was a significant difference in the proportions of patients who achieved a sustained remission when using the lenient criteria (χ^2^ [2, *n* = 96] = 12.63, *p* < 0.01). Overall, of the 580 drug-switch patients, the sustained remission rate during follow-up ranged from 3.1 to 8.4%, depending on the stringency criteria used.

Of the 105 patients who did not meet remission criteria in step-2 drug-switch treatment, 69 (65.7%) relapsed during the follow-up phase and there was no significant difference between the treatment groups (χ^2^ [2, *n* = 105] = .40, *p* = 0.82). However, the proportion of relapses were significantly greater among non-remitted than remitted patients (65.7% vs 29.2%; χ^2^ [1, *n* = 201] = 26.83, p < 0.001).

## Discussion

Our RIAT reanalysis of STAR*D’s step-2 drug-switch patient-level dataset identified several important discrepancies from the original STAR*D published report.^2^

For remission on the HRSD*--the primary outcome--*we found lower rates for each of the step-2 drug-switch treatments compared to those reported by the STAR*D investigators.^2^ This discrepancy resulted from the 147 drug-switch patients that our RIAT reanalysis excluded because these patients failed to meet STAR*D’s stated inclusion for data analysis criteria (see:1, figure 1); and included 44 patients who scored as remitted prior to starting drug-switch treatment despite the STAR*D investigators prespecifying that “*patients who begin a level with HRSD <8 will be excluded from analyses*” ^8, p.130^ (see Supplement 1).

While the drug-switch patients were deemed “*truly resistant*” in step-1 to citalopram therapy by the STAR*D investigators,^1, p.30^ the fact that sertraline, a similar SSRI, was equally ineffective as the other non-SSRI antidepressants suggests that remissions during step-2 therapy were at best only minimally related to the use of other pharmacologically-distinct (and theoretically more effective) step-2 study medications (i.e., bupropion or venlafaxine). This finding comports with a meta-analysis reporting that switching antidepressants after non-response does not improve outcome.^21^

A finding in line with the STAR*D investigators’ summary article was that non-remitted patients who entered follow-up were far more likely to relapse than remitted patients who entered follow-up.^7^

Our RIAT reanalysis found that the overall rate of patients who successfully remitted during step-2 drug-switch therapy, and then sustained this remission during the 12-month follow-up, ranged from only 3.1% to 8.4%. These abysmally low findings corroborate the sustained remission rates reported from STAR*D’s precursor study, the *Texas Medication Algorithm Project* (T-MAP), in which the investigators reported a sustained remission rate between 3.0% to 5.1%.^22^ We note that the STAR*D’s PIs acknowledged in their T-MAP report the following statement: “*Findings from this study reveal remarkably low response and remission rates, and even lower sustained response and remission rates…These findings are especially striking given the fact that treatment was delivered under conditions specifically designed to maximize clinical outcomes*.”^22, p.50^

Although our current RIAT reanalysis found no statistically significant difference in TESI risk rates between the individual step-2 drug-switch treatments, we did identify a significant increase in TESI during drug-switch treatment compared to step-1 citalopram therapy that averaged 30%.

Given the original expressed intention of the STAR*D investigators to achieve remission by administering increasingly more aggressive antidepressant treatments after a failed citalopram trial, one would expect the second, pharmacologically different class of antidepressant to produce a better outcome than the step-2 sertraline treatment (a pharmacologically similar SSRI therapy to citalopram). Furthermore, one might also expect that the relative risk of TESI would decrease (*rather than increase*) during a more aggressive, pharmacologically-distinct antidepressant therapy versus a similar SSRI treatment like sertraline.

In contrast to these expectations, our RIAT reanalysis of the step-2 drug-switch outcomes of effectiveness and TESI rates found an *increase* in the relative risk-to-benefit ratio for all STAR*D step-2 therapies (including that of sertraline). This significant increase in TESI rates might have been due to (1) the switching of medications without first tapering citalopram due to withdrawal issues (however, we would not expect to see a TESI increase in patients switching to another SSRI), (2) the TESI risk profiles of the drug-switch medications relative to citalopram, (3) the loss of hope patients might feel during a second failed drug trial, or (4) other factors,^23^ including persistent side-effect burden.^24^

Severe suicidality emerged in 16 (2.8%) of the step-2 drug-switch patients. STAR*D’s treating clinicians rated these 16 patients as “*Thinks of suicide/death several times a day in depth, or has made specific plans, or attempted suicide*” yet there were no corresponding SAEs for 12 of these patients in either the publication or SAE dataset. While the STAR*D investigators reported no incidences of “Suicide ideation without hospitalization” in their SAE table, the treating clinicians rated that severe-suicidality emerged in 2.8% of patients. In contrast, STAR*D investigators acknowledged severe-suicidality in only four (0.55%) of the treated patients— *an underreporting of 3 out of 4 patients with severe suicidality according to the treating clinicians’ perspective*.

There are several limitations to be considered. For example, there was neither a placebo nor a waitlist control group included in the STAR*D protocol design, which prevented both the STAR*D investigators and our RIAT reanalysis from determining the extent to which these low remission rates occurred from the true pharmacologic effects of the prescribed antidepressant medications *per se*. Similarly, neither our RIAT reanalysis—*nor STAR*D’s*—could evaluate the relationship between the substantial percentage of patients who were prescribed concomitant step-2 psychotropic medications and outcome (e.g., Trazadone: 17.1%, benzodiazepines: 11.8%; and sedative/hypnotics: 16.5%).^2^

In conclusion, our RIAT reanalysis found lower remission rates to acute step-2 drug-switch therapies, compared to the rates reported by the STAR*D investigators.^2^ Furthermore, we found exceedingly low 12-month sustained remission rates during follow-up treatment, a finding which had not previously been reported by the STAR*D investigators.

Similarly, our RIAT reanalysis found an increase in TESI symptoms in all step-2 switch therapies, compared to step-1 treatment with citalopram, suggesting an increase in the risk-to-benefit ratio for the drug-switch therapies. In addition, 12 incidences of severe suicidality noted by the treating clinicians were not reported by the STAR*D investigators.

Thus, neither the original reported findings of the STAR*D step-2 drug-switch outcomes, nor our RIAT reanalysis, could identify any “best” next-step therapy after a failed SSRI trial with citalopram for depression as they were each equally disappointing.

Overall, our RIAT-reanalysis of step-2 drug switch treatments found less optimal effectiveness and more suicidal risk than reported in the original publication, in line with our prior reanalysis of STAR*D’s dataset. Given the world-wide impact of the STAR*D trial in treating MDD, the current RIAT reanalysis continues the process of restoring, with fidelity to the original study protocol, the actual STAR*D results for use in evidence-based treatment. We encourage other independent investigators to examine the original STAR*D patient-level dataset, as well.

## Data Availability

Data used in the preparation of this manuscript were downloaded in January 2025 from the controlled access datasets distributed from the NIMH-supported National Database for Clinical Trials (NDCT).

## Acknowledgements

Data used in the preparation of this manuscript were downloaded in January 2025 from the controlled access datasets distributed from the NIMH-supported National Database for Clinical Trials (NDCT). NDCT is a collaborative informatics system created by the National Institute of Mental Health to provide a national resource to support and accelerate discovery related to clinical trial research in mental health. The content of this publication does not necessarily reflect the views of the RIAT Support Center, NIMH, nor the National Institutes of Health.

## Contributors

HEP, JDA, CX, TK, MP, and IK contributed to the design of the study. CX and TK conducted all of the data analyses. HEP wrote the manuscript with input from CX, JDA, MP, TK, and IK. HEP is responsible for the overall content as the guarantor.

