## Supplementary Appendix for "Restoring STAR*D: A Reanalysis of Drug-Switch Therapy After Failed SSRI Treatment Using Patient-Level Data with Fidelity to the Original STAR*D Research Protocol"

**Table of Contents**

1. Supplement 1: The number of drug-switch patients who were excluded in our reanalysis, yet included in the STAR*D investigators’ step-2 article, and the reasons for their exclusion.
2. Supplement 2: Patient flow chart moving from step-1 citalopram treatment to the step-2 drug-switch therapies.
3. Supplement 3: STAR*D’s system of care designed to maximize remissions while minimizing elapse and dropouts.
4. Supplement 4: Description of the drug-switch medications and their dosing.
5. Supplement 5: Drug-switch patients missing an exit HRSD.
6. Supplement 6: The statistical code used to analyze the STAR*D patient-level dataset.
7. Supplement 7: Demographic and clinical characteristics table.
8. Supplement 8: Comparison of the remissions rates reported in the original publication and those in our RIAT reanalysis.
9. Supplement 9: Calculation of TESI rates for step-2 drug-switch and step-1 citalopram treatments.

**Supplement 1:** **Number of Drug-Switch Patients Excluded from our RIAT**

**Reanalysis, and the Reasons for their Exclusion, yet Included in STAR*D’s Original Publication**

|  | Bup | Sert | Ven | Total |
| --- | --- | --- | --- | --- |
| Scored as Remitted at **ENTRY** into step-2 yet still included in STAR*D’s step-2 analyses | 22 | 8 | 14 | 44 |
| Scored as only mildly depressed (HRSD >7 & <14) at entry into step-1, and therefore excluded from STAR*D’s data analysis, yet still treated in step-1, progressed to step-2, and then included in STAR*D’s step-2 data analyses | 21 | 15 | 25 | 61 |
| Scored as Remitted at entry into step-1 (HRSD ≤ 7), and therefore excluded from STAR*D’s data analysis, yet still treated in step-1 and progressed to step-2 and then included in STAR*D’s step-2 data analyses | 6 | 1 | 4 | 11 |
| Missing baseline HRSD at entry into step-1, and therefore excluded from STAR*D’s data analysis, yet still treated in step-1, and progressed to step-2, and then included in STAR*D’s step-2 data analyses | 12 | 18 | 22 | 52 |
| Number meeting 2 exclusion criterions | 12 | 2 | 7 | 21 |

Bup=Sustained-release Bupropion; Sert= Sertraline; Ven= Extended-release Venlafaxine

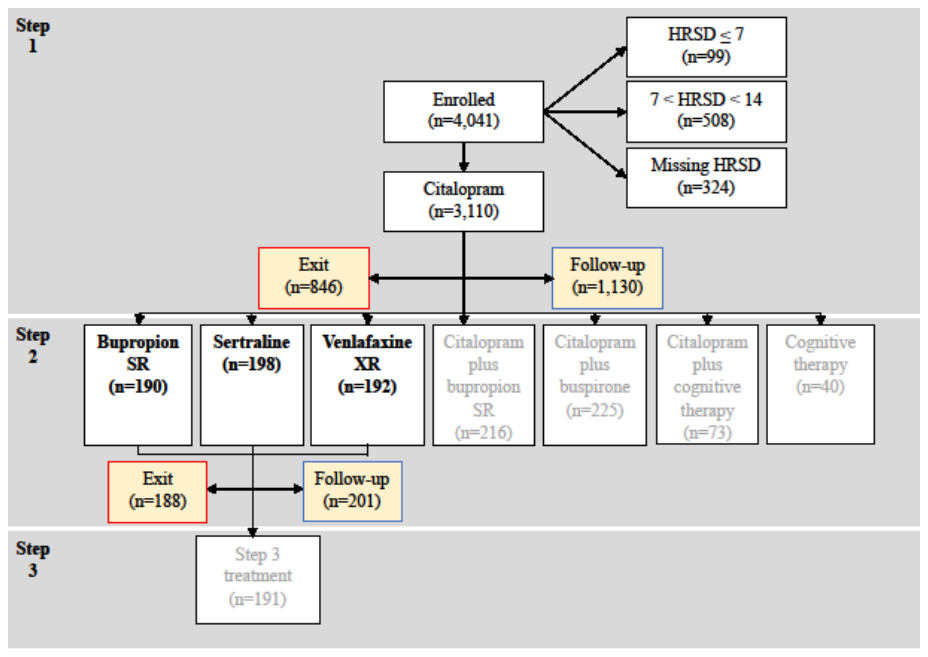

**Supplement 3: Highest Quality of Acute and Continuing-Care to**

**Maximize Remissions While Minimizing Relapse and Dropouts**

| Descriptor | Explanation |
| --- | --- |
| Optimized Sustained Study Participation to Minimize Dropouts | - Promoted patients’ study affiliation via STAR*D-branded brochures, bimonthly newsletters, and an informational video emphasizing STAR*D’s public health significance and the critical role played by patients; - Provided a multistep educational program for patients and families throughout acute-care based on the neurochemical imbalance theory of depression that included “*a glossy visual representation of the brain and neurotransmitters*,” consistently emphasizing that “*depression is a disease, like diabetes or high blood pressure, and has not been caused by something the patient has or has not done. (Depression is an illness, not a personal weakness or character flaw*.) *The educator should emphasize that depression can be treated as effectively as other illnesses*,” and “*explaining the basic principles of mechanism of action*” for the patient’s current antidepressant drug (Patient Education Manual pp. 4–7).; - Used a letter reminder system to alert patients before appointments in those clinics without such systems who had a >15% rate of missed appointments; - Ensured timely follow-up and rescheduling of missed appointments by calling patients on the day of the missed appointment, and again within 24 hours, if there was no response. Patient’s physician sent letter within 48 hours if contact was not established; - Used a letter reminder system for all research outcome assessment calls during acute and continuing-care; - In every clinic visit, the Clinical Research Coordinator (CRC) discussed the research outcomes phone calls with the patient to ensure that the calls were completed on schedule and worked to resolve any problematic issues regarding said calls [Clinical Procedures Manual, page 75]; - Paid patients $25.00 for participating in each telephonic research outcomes assessment; - Permitted patients to re-enter acute and/or continuing-care within four weeks after having dropped out [Clinical Procedures Manual, page 80]; - Recommended one-year of continuing-care for all patients who achieved a satisfactory clinical response with the essential goal of preventing relapse [Clinical Procedures Manual, page 15] and - Permitted continuing-care patients to remain in the study if they moved from the area [Clinical Procedures Manual, page 81]. |
| Acute-Care Visits | Physicians met with patients on entry into each new step to initiate drug treatment with follow-up visits scheduled on weeks 2, 4, 6, 9, 12, with an optional week 14 visit. |
| Measurement-Based Care | Conducted structured evaluations of symptoms and side-effects at each visit and included a centralized treatment monitoring and physician feedback system to ensure consistent implementation of optimal care across research sites. |
| Aggressive Medication Dosing | Provided aggressive medication dosing with a fully adequate dose for a sufficient duration to “*ensure that the likelihood of achieving remission was maximized and that those who did not reach remission were truly resistant to the medication*”. |
| Liberal Prescribing of Non-Study Medications | Physicians had great leeway in prescribing non-study medications to treat comorbid symptoms resulting in:   - 17.2% taking Trazodone for sleep; - 11.9% taking an anti-anxiety medication; - 16.7% taking either a sedative or hypnotic medication; and - An undisclosed percent taking medications to address side-effects. |
| Continuing-Care Visits | Patients saw their physician every 2 months and continued taking their treatment medication(s) at the same doses but their physicians were allowed to make any psychotherapy, medication, and/or medication dose changes to maximize the likelihood of maintaining patients’ remission status. Additional continuing-care visits were scheduled when patients began to experience a return of depressive symptoms and/or intolerable side-effects [Clinical Procedures Manual. page 78]. |
| Clinical Research Coordinator (CRC) | Each site had a CRC who:   - Saw patients before each visit administering multiple measures to them including the QIDS-SR during each acute-care visit; - Assisted physicians in protocol implementation; and - Provided patients support and encouragement in protocol implementation. |
| Treatment Designed to Enhance Subject Retention | Treatment was designed to minimize drop-outs and/or non-compliance including:   - Open label prescribing during acute and continuing-care with no placebo control condition during any study phase; - Patients chose their acceptable treatment assignments for steps two and three to eliminate any concerns they might have about receiving an unacceptable assignment. - During each step, patients could enroll immediately into the next step if they had intolerable side-effects or had maximized their current medication(s)’ dosing without achieving a remission; and - During any step, patients could enter continuing-care directly on their current medication(s) if they were treatment responders even if they had not achieved remission. This was done to minimize responders from dropping out in order to avoid having to discontinue their current medication(s) and start a new drug regimen. |

O’Neal, B., & Biggs, M. (2001). STAR*D patient education manual. Retrieved July 5, 2010, from http://www.edc.pitt.edu/stard/public/study_manuals.html

Trivedi MH, Stegman D, Rush AJ, Wisniewski SR, Nierenberg AA: STAR*D clinical procedures manual. July 31, 2002. www.edc.pitt. edu/stard/public/study_manuals.html

**Supplement 4:**

**Description of Drug-Switch Treatments**

Citalopram was discontinued without a tapering at the initiation of each step-2 switch treatment. STAR*D investigators chose pharmacologically-distinct switch medications. The three drug-switch treatments were:

- Sustained-release bupropion (Wellbutrin SR), an “out-of-class” agent whose neurochemical action mechanisms are unknown; other than that, it does not inhibit serotonin reuptake and is believed to produce antidepressant effects by blocking the reuptake of dopamine and norepinephrine. The daily dose of sustained-release bupropion was 150 mg for week 1, 200 mg from day 8 to 27, 300 mg from day 28 to 41, and 400 mg from day 42 onward.
- Sertraline (Zoloft), an SSRI with the same pharmacological profile as citalopram. Sertraline was started at a daily dose of 50 mg and increased to 100 mg at day 8, to 150 mg at day 28, and to 200 mg at day 63 and onward.
- Extended-release venlafaxine (Effexor), a “dual-action” agent that inhibits the reuptake of both serotonin and norepinephrine. The starting daily dose of extended-release venlafaxine was 37.5 mg for week 1 and increased to 75 mg from day 8 to 14, to 150 mg from day 15 to 27, to 225 mg from day 28 to 41, to 300 mg from day 42 to 62, and to 375 mg from day 63 onward.

**Supplement 5: Number and Percent of Drug-Switch**

**Patients Missing Exit HRSD**

|  | **#/(%) with Missing Exit HRSD** |
| --- | --- |
| Bupropion (N=190) | 58 (30.5%) |
| Sertraline (N=198) | 56 (28.3%) |
| Venlafaxine (N=192) | 56 (29.2%) |
| Total (N=580) | 170 (29.3%) |

**Supplement 6:**

**Definitions of Acute and Follow-up Care Outcomes and the Statistical**

**Code Used to Analyze the Dataset**

Response was defined as a ≥ 50% reduction in HRSD between baseline level 2 HRSD (defined as the exit HRSD from level 1, or if they are missing exit HRSD from level, the HRSD mapped from QIDS at the last observation of level 1) and the exit for level 2 HRSD (defined as the last HRSD for level 2, or if they are missing HRSD from that level, the HRSD mapped from the QIDS at the last observation of level 2).

We also calculated mean change in HRSD score between baseline level 2 HRSD (defined as the exit HRSD from the previous level, in this case, level 1, or if they are missing exit HRSD from level, the HRSD mapped from QIDS at the last observation of the previous level) and the exit for level 2 HRSD (defined as the last HRSD for level 2, or if they are missing HRSD from that level, the HRSD mapped from the QIDS at the last observation of level 2).

```
####################################################################################################
####################			READING IN RAW HRSD DATA				####################
####################################################################################################

### read in p01 version hrsd Data
hrsdP01Header		<-	read.table("InputFolder/Data Files/NIMH p01 Nov 2019 Data/hrsd01.txt", nrows = 1, header = FALSE, stringsAsFactors = FALSE)		#Extract current file header
hrsdP01Raw 			<-	read.table("InputFolder/Data Files/NIMH p01 Nov 2019 Data/hrsd01.txt", skip = 2, header = FALSE, fill = T)						#Extract current file data
colnames(hrsdP01Raw ) 	<- unlist(hrsdP01Header)										#Merge file Fheader with data

### read in ivra data
ivraP01Header		<-	read.table("InputFolder/Data Files/NIMH p01 Nov 2019 Data/ivra01.txt", nrows = 1, header = FALSE, stringsAsFactors = FALSE)		#Extract current file header
ivraP01Raw 			<-	read.table("InputFolder/Data Files/NIMH p01 Nov 2019 Data/ivra01.txt", skip = 2, header = FALSE, fill = T)						#Extract current file data
colnames(ivraP01Raw ) 	<- unlist(ivraP01Header)										#Merge file header with data

### read in QIDS data
qidsP01Header		<-	read.table("InputFolder/Data Files/NIMH p01 Nov 2019 Data/qids01.txt", nrows = 1, header = FALSE, stringsAsFactors = FALSE)		#Extract current file header
qidsP01Raw 			<-	read.table("InputFolder/Data Files/NIMH p01 Nov 2019 Data/qids01.txt", skip = 2, header = FALSE, fill = T)						#Extract current file data
colnames(qidsP01Raw ) 	<- unlist(qidsP01Header)										#Merge file header with data

#########################################################################################
####################		TASK 1: RESCORING VARIABLES		###################
#########################################################################################

##################  SCORING HRSD-17		################
# Since hdtot_r variable is missing for all levels except enrollment, we need to manually rescore HRSD
#

## Find out how many have hdtot_r
table(is.na(hrsdP01Raw$hdtot_r))			# Turns out only 4039 scored hdtot_r's available

## Find which levels have legitimate hdtot_r	##
table(hrsdP01Raw[!is.na(hrsdP01Raw$hdtot_r),"level"])		# turns out only enrollment has the HRSD sum score

## Look only at enrollment data since they have a valid hdtot_r	##
table(hrsdP01Raw[hrsdP01Raw$level =="Enrollment",]$hdtot_r , useNA="always")

## Rescoring by adding symptom vars	##

hrsdSymptomVars	<- c("hsoin", "hmnin", "hemin", "hmdsd", "hpanx", "hinsg", "happt", "hwl", "hsanx", "hhypc", "hvwsf", "hsuic", "hintr", "hengy", "hslow", "hagit", "hsex")

hrsdP01Raw$hrsdtot_sum_NAasNA			<- rowSums(hrsdP01Raw[,c(hrsdSymptomVars)])			#This version treats NA's as zeros

### Checking IVRA -- Action_call shows when people are moved to followup OR to new treatment	###
with(ivraP01Raw, table(action_call, txassign))

##################  Scoring QIDS data since Followup is lacking qstot		################
# Since followup QIDs is laking a qstot totalscore, we need to manually score QIDS data
#

# Sleep variables
qidsP01Raw $maxSleep 	<- apply(qidsP01Raw [, c("vsoin", "vmnin", "vhysm", "vemin")], MARGIN = 1, max, na.rm = T)

# Add that result to the maximum score of the weight/appetite variables (SAPDC, SAPIN, SWTDC, SWTIN)
qidsP01Raw $maxAppetite 	<- apply(qidsP01Raw [, c("vapdc", "vapin", "vwtdc", "vwtin")], MARGIN = 1, max, na.rm = T)

# Then add that sum to the maximum of the two motor variables (SAGIT, SSLOW)
qidsP01Raw $maxMotor 	<- apply(qidsP01Raw [, c("vagit", "vslow")], MARGIN = 1, max, na.rm = T)

# And finally add to that sum the sum of the rest of the scores (SMDSD, SCNTR, SVWSF, SSUIC, SINTR, SENGY)

qidsP01Raw $sumOther 	<- rowSums(qidsP01Raw [, c("vmdsd", "vcntr", "vvwsf", "vsuic", "vintr", "vengy")])
qidsP01Raw $SR_TOTAL <- rowSums(qidsP01Raw [, c("maxSleep", "maxAppetite", "maxMotor", "sumOther")])

########################################################################
##############		Function to remap QIDS		##################
########################################################################

# In order to impute missing HRSD scores using QIDS scores, as per methods section of BMJ Open paper

remapHRSD_QIDS	<- function(x){
	y <- NA
	x <- as.character(x)
	switch(x,
		'1' =  {y <- 1.5},
		'2' =  {y <- 3},
		'3' =  {y <- 4},
		'4' =  {y <- 5.5},
		'5' =  {y <- 7},
		'6' =  {y <- 8},
		'7' =  {y <- 9.5},
		'8' =  {y <- 11},
		'9' =  {y <- 12},
		'10' =  {y <- 13},
		'11' =  {y <- 14.5},
		'12' =  {y <- 16},
		'13' =  {y <- 17},
		'14' =  {y <- 18.5},
		'15' =  {y <- 18.5},
		'16' =  {y <- 20},
		'17' =  {y <- 21.5},
		'18' =  {y <- 23},
		'19' =  {y <- 24},
		'20' =  {y <- 25},
		'21' =  {y <- 26.5},
		'22' =  {y <- 28},
		'23' =  {y <- 29},
		'24' =  {y <- 30.5},
		'25' =  {y <- 32},
		'26' =  {y <- 34},
		'27' =  {y <- 44})
	return(y)
	}

##########################################################################################################
##################		DEFINE WINDOWS FOR FOLLOW UP HRSD		#####################################
##########################################################################################################

###	Define month windows for QIDS	###
#
# Follow up HRSD and QIDS
# Find out if they have a month corresponding HRSD. If they do not, then replace it with a QIDS from the specified window (SR_TOTAL). (Have as two outcomes)
#
# 4 time points:
# Month 3 - Window 63-119
# Month 6 - Window 154-210
# Month 9 - Window 245-301
# Month 12 - Window 336-392
#
# If they have observation not in windows, count it as missing
#
# For month on hrsd variable, use month variable (which describes monthin follow up, -2 for those who do not have followup)
# For days, use days_baseline from QIDS
#
######################################

	qidsWindowMonth3_lo	<- 63
	qidsWindowMonth3_hi	<- 119
	qidsWindowMonth6_lo	<- 154
	qidsWindowMonth6_hi	<- 210
	qidsWindowMonth9_lo	<- 245
	qidsWindowMonth9_hi	<- 301
	qidsWindowMonth12_lo	<- 336
	qidsWindowMonth12_hi	<- 392

	qidsWindowMonth3_mid	<- 91
	qidsWindowMonth6_mid	<- 182
	qidsWindowMonth9_mid	<- 273
	qidsWindowMonth12_mid	<- 364

###########################################################################################################################
####################		TASK 2: Pulling out HRSD scores, calculating remissions				###################
############################################################################################################################

# First define all subjectID's
allHRSD_subjectID <- unique(hrsdP01Raw$src_subject_id)

# Create an output file for HRSD

stardOutputs	<- data.frame(src_subject_id = allHRSD_subjectID,

							meetInclusion	= NA,

							# IVRA level entry variables

							maxlevel_ivra= NA,

							level1_ivra_days_enter = NA,
							level2_ivra_days_enter = NA,
							level2A_ivra_days_enter = NA,
							level3_ivra_days_enter = NA,
							level4_ivra_days_enter = NA,
							fu_ivra_days_enter   = NA,

							level1_ivra_entered = NA,
							level2_ivra_entered = NA,
							level2A_ivra_entered = NA,
							level3_ivra_entered = NA,
							level4_ivra_entered = NA,
							fu_ivra_entered  = NA,

							# Level 1 variables

							level1_first_hrsd_day = NA,
							level1_first_hrsd_sum = NA,
							level1_last_hrsd_day = NA,
							level1_last_hrsd_sum = NA,

							level1_nHRSD	= NA,

							level1_last_qids_day 	= NA,
							level1_last_qids_qstot = NA,

							level1_tx = NA,
							level1_remitted = NA,

							level1_remitted_imputed = NA,
							level1_first_hrsd_imputedQIDS	= NA,
							level1_last_hrsd_imputedQIDS = NA,

							level1_responded = NA,
							level1_hrsdchange = NA,
							level1_responded_imputed = NA,
							level1_hrsdchange_imputed = NA,

							# Level 2 variables

							level2_hasHRSDData = NA,
							level2_last_hrsd_day = NA,
							level2_last_hrsd_sum= NA,
							level2_last_qids_day 	= NA,
							level2_last_qids_qstot = NA,

							level2_tx = NA,
							level2_remitted = NA,

							level2_remitted_imputed = NA,
							level2_last_hrsd_imputedQIDS = NA,
							level2_responded = NA,
							level2_hrsdchange = NA,
							level2_responded_imputed = NA,
							level2_hrsdchange_imputed = NA,

							# Level 2A variables

							level2A_hasHRSDData = NA,
							level2A_last_hrsd_day = NA,
							level2A_last_hrsd_sum= NA,
							level2A_last_qids_day 	= NA,
							level2A_last_qids_qstot = NA,

							level2A_tx = NA,
							level2A_remitted = NA,

							level2A_remitted_imputed = NA,
							level2A_last_hrsd_imputedQIDS = NA,
							level2A_responded = NA,
							level2A_hrsdchange = NA,
							level2A_responded_imputed = NA,
							level2A_hrsdchange_imputed = NA,

							# Level 3 variables

							level3_hasHRSDData = NA,
							level3_last_hrsd_day = NA,
							level3_last_hrsd_sum= NA,

							level3_last_qids_day 	= NA,
							level3_last_qids_qstot = NA,

							level3_tx = NA,
							level3_remitted = NA,

							level3_remitted_imputed = NA,
							level3_last_hrsd_imputedQIDS = NA,
							level3_responded = NA,
							level3_hrsdchange = NA,
							level3_responded_imputed = NA,
							level3_hrsdchange_imputed = NA,

							# Level 4 variables

							level4_hasHRSDData = NA,
							level4_last_hrsd_day = NA,
							level4_last_hrsd_sum= NA,
							level4_last_qids_day 	= NA,
							level4_last_qids_qstot = NA,

							level4_tx = NA,
							level4_remitted = NA,

							level4_remitted_imputed = NA,
							level4_last_hrsd_imputedQIDS = NA,

							level4_responded = NA,
							level4_hrsdchange = NA,
							level4_responded_imputed = NA,
							level4_hrsdchange_imputed = NA,

							# Followup

							fu_hasHRSDData = NA,
							fu_hrsd_month3 = NA,
							fu_hrsd_month6 = NA,
							fu_hrsd_month9 = NA,
							fu_hrsd_month12 = NA,
							fu_qids_month3 = NA,
							fu_qids_month6 = NA,
							fu_qids_month9 = NA,
							fu_qids_month12 = NA,
							fu_qids_month3_date = NA,
							fu_qids_month6_date = NA,
							fu_qids_month9_date = NA,
							fu_qids_month12_date = NA,

							fu_noObservations = NA,

							fu_hrsdImputed_month3 = NA,
							fu_hrsdImputed_month6 = NA,
							fu_hrsdImputed_month9 = NA,
							fu_hrsdImputed_month12 = NA,

							fu_remissionbeforeFU	= NA,
							fu_remissionbeforeFU_imputed = NA,

							fu_relapse = NA,
							fu_susRemission_full = NA,
							fu_susRemission_weak = NA,

							flagged = NA)

# Now loop through subjectID's in Level1	#
for(i in 1:length(allHRSD_subjectID )){

	# Pull out current subject ID
	currentID				<- allHRSD_subjectID[i]

	#Pull out Level1 data, for current subject

	currentSubjectHRSD_all		<- hrsdP01Raw[hrsdP01Raw$src_subject_id == currentID,]

	currentSubjectHRSD_Level1		<- hrsdP01Raw[hrsdP01Raw$level == "Level 1" & hrsdP01Raw$src_subject_id == currentID,]
	currentSubjectHRSD_Level2		<- hrsdP01Raw[hrsdP01Raw$level == "Level 2" & hrsdP01Raw$src_subject_id == currentID,]
	currentSubjectHRSD_Level2A	<- hrsdP01Raw[hrsdP01Raw$level == "Level 2A" & hrsdP01Raw$src_subject_id == currentID,]
	currentSubjectHRSD_Level3		<- hrsdP01Raw[hrsdP01Raw$level == "Level 3" & hrsdP01Raw$src_subject_id == currentID,]
	currentSubjectHRSD_Level4		<- hrsdP01Raw[hrsdP01Raw$level == "Level 4" & hrsdP01Raw$src_subject_id == currentID,]

	currentSubjectHRSD_fu	<- hrsdP01Raw[hrsdP01Raw$level == "Follow up" & hrsdP01Raw$src_subject_id == currentID,]

	########## 	IVRA LEVELS		##########

	#Pull out the ivra data for participant
	currentSubject_ivra		<- ivraP01Raw[ivraP01Raw$src_subject_id == currentID,]

	#Pull out corresponding day someone entered a level
	level1_ivra_days_enter  <-  unique(currentSubject_ivra[currentSubject_ivra$level == "Level 1" & currentSubject_ivra$action_call == 1, "days_baseline"])
	level2_ivra_days_enter  <-  unique(currentSubject_ivra[currentSubject_ivra$level == "Level 2" & currentSubject_ivra$action_call == 1 , "days_baseline"])
	level2A_ivra_days_enter <-  unique(currentSubject_ivra[currentSubject_ivra$level == "Level 2A" & currentSubject_ivra$action_call == 1, "days_baseline"])
	level3_ivra_days_enter  <-  unique(currentSubject_ivra[currentSubject_ivra$level == "Level 3" & currentSubject_ivra$action_call == 1, "days_baseline"])
	level4_ivra_days_enter  <-  unique(currentSubject_ivra[currentSubject_ivra$level == "Level 4" & currentSubject_ivra$action_call == 1, "days_baseline"])
	fu_ivra_days_enter  <-  unique(currentSubject_ivra[currentSubject_ivra$level == "Follow up" & currentSubject_ivra$action_call == 2, "days_baseline"])

	# Clean up IVRA for people who did not enter a level
	level1_ivra_days_enter   <- ifelse(length(level1_ivra_days_enter) != 0, level1_ivra_days_enter , NA)
	level2_ivra_days_enter   <- ifelse(length(level2_ivra_days_enter) != 0, level2_ivra_days_enter , NA)
	level2A_ivra_days_enter   <- ifelse(length(level2A_ivra_days_enter) != 0, level2A_ivra_days_enter , NA)
	level3_ivra_days_enter   <- ifelse(length(level3_ivra_days_enter) != 0, level3_ivra_days_enter , NA)
	level4_ivra_days_enter   <- ifelse(length(level4_ivra_days_enter) != 0, level4_ivra_days_enter , NA)
	fu_ivra_days_enter   	<- ifelse(length(level4_ivra_days_enter) != 0, fu_ivra_days_enter , NA)

	# binary to specify if they entered a level or not
	level1_ivra_entered  <- ifelse(nrow(currentSubject_ivra[currentSubject_ivra$action_call == 1 & currentSubject_ivra$level == "Level 1",]) > 0, 1,0)
	level2_ivra_entered  <- ifelse(nrow(currentSubject_ivra[currentSubject_ivra$action_call == 1 & currentSubject_ivra$level == "Level 2",]) > 0, 1,0)
	level2A_ivra_entered  <- ifelse(nrow(currentSubject_ivra[currentSubject_ivra$action_call == 1 & currentSubject_ivra$level == "Level 2A",]) > 0, 1,0)
	level3_ivra_entered  <- ifelse(nrow(currentSubject_ivra[currentSubject_ivra$action_call == 1 & currentSubject_ivra$level == "Level 3",]) > 0, 1,0)
	level4_ivra_entered  <- ifelse(nrow(currentSubject_ivra[currentSubject_ivra$action_call == 1 & currentSubject_ivra$level == "Level 4",]) > 0, 1,0)
	fu_ivra_entered  		<- ifelse(nrow(currentSubject_ivra[currentSubject_ivra$action_call == 2 & currentSubject_ivra$level == "Follow up",]) > 0, 1,0)

	#Calculate max level, we work backyards incase someone has skipped a level
	maxlevel_ivra		<- ifelse(level4_ivra_entered == 1 , "Level 4",
					ifelse(level3_ivra_entered == 1 , "Level 3",
					ifelse(level2A_ivra_entered == 1 , "Level 2A",
					ifelse(level2_ivra_entered == 1 , "Level 2",
					ifelse(level1_ivra_entered == 1 , "Level 1", NA)))))

	### save IVRA outputs	###
	stardOutputs[stardOutputs$src_subject_id == currentID, "maxlevel_ivra"] <- maxlevel_ivra

	stardOutputs[stardOutputs$src_subject_id == currentID, "level1_ivra_days_enter"] 	<- level1_ivra_days_enter
	stardOutputs[stardOutputs$src_subject_id == currentID, "level2_ivra_days_enter"] 	<- level2_ivra_days_enter
	stardOutputs[stardOutputs$src_subject_id == currentID, "level2A_ivra_days_enter"] 	<- level2A_ivra_days_enter
	stardOutputs[stardOutputs$src_subject_id == currentID, "level3_ivra_days_enter"] 	<- level3_ivra_days_enter
	stardOutputs[stardOutputs$src_subject_id == currentID, "level4_ivra_days_enter"] 	<- level4_ivra_days_enter
	stardOutputs[stardOutputs$src_subject_id == currentID, "fu_ivra_days_enter"] 		<- fu_ivra_days_enter

	stardOutputs[stardOutputs$src_subject_id == currentID, "level1_ivra_entered"] <- level1_ivra_entered
	stardOutputs[stardOutputs$src_subject_id == currentID, "level2_ivra_entered"] <- level2_ivra_entered
	stardOutputs[stardOutputs$src_subject_id == currentID, "level2A_ivra_entered"] <- level2A_ivra_entered
	stardOutputs[stardOutputs$src_subject_id == currentID, "level3_ivra_entered"] <- level3_ivra_entered
	stardOutputs[stardOutputs$src_subject_id == currentID, "level4_ivra_entered"] <- level4_ivra_entered
	stardOutputs[stardOutputs$src_subject_id == currentID, "fu_ivra_entered"] 	<- fu_ivra_entered

	##########	 HRSD DATA 		##########

	### HRSD Level 1 ###

	#Count how many HRSD Rows in level 1
	level1_nHRSD 	<- nrow(unique(currentSubjectHRSD_Level1))

	#Select only rows where days_baseline <= 5
	currentSubjectHRSD_L1_firstLevel15	<- unique(currentSubjectHRSD_Level1[currentSubjectHRSD_Level1$days_baseline <=5 ,])

	#For subjects with multiple before days_baseline <=5, collapse both dates so we can see
	level1_multiple_daysbaseline_5days	<- paste(currentSubjectHRSD_L1_firstLevel15$days_baseline, collapse = "-")
	level1_multiple_hrsd_5days		<- paste(currentSubjectHRSD_L1_firstLevel15$hrsdtot_sum_NAasNA, collapse = "-")

	#We want the earliest one, so pick the earliest row (if applicable)
	currentSubject_L1_earliestHRSD	<- 	unique(currentSubjectHRSD_L1_firstLevel15[order(currentSubjectHRSD_L1_firstLevel15$days_baseline),])[1,]

	level1_first_hrsd_day <- currentSubject_L1_earliestHRSD$days_baseline
	level1_first_hrsd_sum <- currentSubject_L1_earliestHRSD$hrsdtot_sum_NAasNA

	#Find the latest HRSD day
	level1_latestHRSD_day	<- unique(max(currentSubjectHRSD_Level1$days_baseline))

	# Pull out row matching lastLevel1 day, make sure it is only unique. If multple rows then pull out the firstLevel1.
	level1_last_hrsd_rows	<- unique(currentSubjectHRSD_Level1[currentSubjectHRSD_Level1$days_baseline == 	level1_latestHRSD_day & !is.na(currentSubjectHRSD_Level1$days_baseline ) & !is.na(currentSubjectHRSD_Level1$hrsdtot_sum_NAasNA),])[1,]

	# Require lastLevel1 HRSD to be over 10 days
	level1_last_hrsd_rows	<- level1_last_hrsd_rows[level1_last_hrsd_rows$days_baseline > 10,]

	if (nrow(level1_last_hrsd_rows) == 0 ){
		level1_last_hrsd_day		<- NA
		level1_last_hrsd_sum	<- NA
	}else {
		level1_last_hrsd_day			<- level1_last_hrsd_rows$days_baseline
		level1_last_hrsd_sum			<- level1_last_hrsd_rows$hrsdtot_sum_NAasNA
	}

	# Check if they meet inclusion criteria
	meetInclusion	<-	ifelse(is.na(currentSubject_L1_earliestHRSD$hrsdtot_sum_NAasNA) | currentSubject_L1_earliestHRSD$hrsdtot_sum_NAasNA <  14,0,1)

	# Check if they remitted in level 1
	level1_remitted <- ifelse(meetInclusion == 1 & !is.na(level1_last_hrsd_sum) & level1_last_hrsd_sum<= 7, 1,
					ifelse(is.na(level1_last_hrsd_sum), NA, 0))
	# Find level 1 treatment condition
	level1_tx <- ivraP01Raw[ivraP01Raw$src_subject_id == currentID & ivraP01Raw$level == "Level 1" & ivraP01Raw$txassign != "", "txassign"]

	# Calculate HRSD change
	level1_hrsdchange	<- level1_last_hrsd_sum - level1_first_hrsd_sum

	# Calculate response, defined as 50% improvement from last to first
	level1_responded	<- ifelse((-level1_hrsdchange / level1_first_hrsd_sum) >= 0.50, 1, 0)

	#Now save this to HRSD Output File
	stardOutputs[stardOutputs$src_subject_id == currentID, "level1_first_hrsd_day"] 			<- level1_first_hrsd_day
	stardOutputs[stardOutputs$src_subject_id == currentID, "level1_first_hrsd_sum"] 			<- level1_first_hrsd_sum

	stardOutputs[stardOutputs$src_subject_id == currentID, "level1_last_hrsd_sum"] 			<- level1_last_hrsd_sum
	stardOutputs[stardOutputs$src_subject_id == currentID, "level1_last_hrsd_day"] 			<-  level1_last_hrsd_day
	stardOutputs[stardOutputs$src_subject_id == currentID, "level1_nHRSD"] 				<- level1_nHRSD
	stardOutputs[stardOutputs$src_subject_id == currentID, "level1_remitted"] 			<- level1_remitted

	stardOutputs[stardOutputs$src_subject_id == currentID, "level1_hrsdchange"] 			<- level1_hrsdchange
	stardOutputs[stardOutputs$src_subject_id == currentID, "level1_responded"] 			<- level1_responded

	stardOutputs[stardOutputs$src_subject_id == currentID, "meetInclusion"] 				<- meetInclusion
	stardOutputs[stardOutputs$src_subject_id == currentID, "level1_tx"] 				<- level1_tx

	### Now do Level 2 HRSD ###

	if(nrow(currentSubjectHRSD_Level2) == 0){		# If they do not have level 2, specify everything as NA

		level2_hasHRSDData	<- 0
		level2_last_hrsd_sum	<-  NA
		level2_last_hrsd_day	<-  NA
		level2_remitted 		<-  NA
		level2_responded		<- NA
		level2_hrsdchange 		<- NA

	}else {			#Otherwise calculate level 2 data

		level2_hasHRSDData	<- 1

		#Find the latest HRSD day
		level2_last_hrsd_day	<- unique(max(currentSubjectHRSD_Level2$days_baseline))

		# Pull out row matching last day, make sure it is only unique. If multple rows then pull out the first.
		level2_last_hrsd_rows	<- unique(currentSubjectHRSD_Level2[currentSubjectHRSD_Level2$days_baseline == level2_last_hrsd_day& !is.na(currentSubjectHRSD_Level2$days_baseline ) ,])[1,]

		# Pull out last level 2 hrsd sum
		level2_last_hrsd_sum<- level2_last_hrsd_rows$ hrsdtot_sum_NAasNA

		##Calculate level 2 remitted: meetInclusion = 1, did not remit in level 1, and level 2 last HRSD <= 7
		level2_remitted <- ifelse(meetInclusion == 1  & level2_last_hrsd_sum<= 7, 1, 0)

		# Calculate HRSD change in level 2 -- using last level 1 HRSD as level 2 baseline
		level2_hrsdchange	<- level2_last_hrsd_sum - level1_last_hrsd_sum

		# Calculate response, defined as 50% improvement from last level 1 HRSD
		level2_responded	<- ifelse((-level2_hrsdchange / level1_last_hrsd_sum) >= 0.50, 1, 0)

	}

	# Pull out level2 treatment
	level2_tx <- ivraP01Raw[ivraP01Raw$src_subject_id == currentID & ivraP01Raw$level == "Level 2" & ivraP01Raw$txassign != "", "txassign"]

	# Convert level 2 treatment to NA for people who did not get assigned
	level2_tx <- ifelse(length(level2_tx) != 0, level2_tx, NA)

	## Now saving it to output
	stardOutputs[stardOutputs$src_subject_id == currentID, "level2_hasHRSDData"] 			<- level2_hasHRSDData
	stardOutputs[stardOutputs$src_subject_id == currentID, "level2_last_hrsd_sum"] 			<- level2_last_hrsd_sum
	stardOutputs[stardOutputs$src_subject_id == currentID, "level2_last_hrsd_day"] 			<- level2_last_hrsd_day
	stardOutputs[stardOutputs$src_subject_id == currentID, "level2_remitted"] 				<- level2_remitted
	stardOutputs[stardOutputs$src_subject_id == currentID, "level2_tx"] 					<- level2_tx

	stardOutputs[stardOutputs$src_subject_id == currentID, "level2_hrsdchange"] 				<- level2_hrsdchange
	stardOutputs[stardOutputs$src_subject_id == currentID, "level2_responded"] 				<- level2_responded

	### Now do Level 2 A HRSD ###

	if(nrow(currentSubjectHRSD_Level2A) == 0){		# If they do not have Level 2 A, specify everything as NA
		level2A_hasHRSDData	<- 0
		level2A_last_hrsd_sum	<-  NA
		level2A_last_hrsd_day	<-  NA
		level2A_remitted 		<-  NA

		level2A_responded		<- NA
		level2A_hrsdchange 		<- NA

	}else {			#Otherwise calculate Level 2 A data

		level2A_hasHRSDData	<- 1

		#Find the latest HRSD day
		level2A_last_hrsd_day	<- unique(max(currentSubjectHRSD_Level2A$days_baseline))

		# Pull out row matching last day, make sure it is only unique. If multple rows then pull out the first.
		level2A_last_hrsd_rows	<- unique(currentSubjectHRSD_Level2A[currentSubjectHRSD_Level2A$days_baseline == level2A_last_hrsd_day& !is.na(currentSubjectHRSD_Level2A$days_baseline ) ,])[1,]

		# Pull out last Level 2 A hrsd sum
		level2A_last_hrsd_sum <- level2A_last_hrsd_rows$ hrsdtot_sum_NAasNA

		##Calculate Level 2 A remitted: meetInclusion = 1, did not remit in level 1, and Level 2 A last HRSD <= 7
		level2A_remitted <- ifelse(meetInclusion == 1 & level2A_last_hrsd_sum <= 7, 1, 0)

		# Calculate HRSD change from last level 2 HRSD
		level2A_hrsdchange	<- level2A_last_hrsd_sum - level2_last_hrsd_sum

		# Calculate response, defined as 50% improvement from last level 2 HRSD
		level2A_responded	<- ifelse((-level2A_hrsdchange / level2_last_hrsd_sum) >= 0.50, 1, 0)

	}

	# Pull out level2A treatment
	level2A_tx <- ivraP01Raw[ivraP01Raw$src_subject_id == currentID & ivraP01Raw$level == "Level 2A" & ivraP01Raw$txassign != "", "txassign"]

	# Convert Level 2 A treatment to NA for people who did not get assigned
	level2A_tx <- ifelse(length(level2A_tx) != 0, level2A_tx, NA)

	## Now saving it to output
	stardOutputs[stardOutputs$src_subject_id == currentID, "level2A_hasHRSDData"] 			<- level2A_hasHRSDData
	stardOutputs[stardOutputs$src_subject_id == currentID, "level2A_last_hrsd_sum"] 			<- level2A_last_hrsd_sum
	stardOutputs[stardOutputs$src_subject_id == currentID, "level2A_last_hrsd_day"] 			<- level2A_last_hrsd_day
	stardOutputs[stardOutputs$src_subject_id == currentID, "level2A_remitted"] 			<- level2A_remitted
	stardOutputs[stardOutputs$src_subject_id == currentID, "level2A_tx"] 				<- level2A_tx

	stardOutputs[stardOutputs$src_subject_id == currentID, "level2A_hrsdchange"] 			<- level2A_hrsdchange
	stardOutputs[stardOutputs$src_subject_id == currentID, "level2A_responded"] 			<- level2A_responded

	### Now do Level 3 HRSD ###

	if(nrow(currentSubjectHRSD_Level3) == 0){		# If they do not have Level 3, specify everything as NA
		level3_hasHRSDData	<- 0
		level3_last_hrsd_sum	<-  NA
		level3_last_hrsd_day	<-  NA
		level3_remitted 		<-  NA
		level3_hrsdchange		<-  NA
		level3_responded		<-  NA
		level3_entry_hrsd_sum 	<-  NA

	}else {			#Otherwise calculate Level 3 data

		level3_hasHRSDData	<- 1

		#Find the latest HRSD day
		level3_last_hrsd_day	<- unique(max(currentSubjectHRSD_Level3$days_baseline))

		# Pull out row matching last day, make sure it is only unique. If multple rows then pull out the first.
		level3_last_hrsd_rows	<- unique(currentSubjectHRSD_Level3[currentSubjectHRSD_Level3$days_baseline == level3_last_hrsd_day& !is.na(currentSubjectHRSD_Level3$days_baseline ) ,])[1,]

		# Pull out last Level 3 hrsd sum
		level3_last_hrsd_sum <- level3_last_hrsd_rows$ hrsdtot_sum_NAasNA

		##Calculate Level 3 remitted: meetInclusion = 1, did not remit in level 1, and Level 3 last HRSD <= 7
		level3_remitted <- ifelse(meetInclusion == 1 & level3_last_hrsd_sum <= 7, 1, 0)

		## Find level 3 entry HRSD depending on if they entered level 2A or not
		level3_entry_hrsd_sum	<- ifelse(level2A_hasHRSDData == 1, level2A_last_hrsd_sum , level2_last_hrsd_sum )

		# Calculate HRSD change
		level3_hrsdchange	<- level3_last_hrsd_sum - level3_entry_hrsd_sum

		# Calculate response, defined as 50% improvement from last to first
		level3_responded	<- ifelse((-level3_hrsdchange / level3_entry_hrsd_sum ) >= 0.50, 1, 0)

	}

	# Pull out level3 treatment
	level3_tx <- ivraP01Raw[ivraP01Raw$src_subject_id == currentID & ivraP01Raw$level == "Level 3" & ivraP01Raw$txassign != "", "txassign"]

	# Convert Level 3 treatment to NA for people who did not get assigned
	level3_tx <- ifelse(length(level3_tx) != 0, level3_tx, NA)

	## Now saving it to output
	stardOutputs[stardOutputs$src_subject_id == currentID, "level3_hasHRSDData"] 				<- level3_hasHRSDData
	stardOutputs[stardOutputs$src_subject_id == currentID, "level3_last_hrsd_sum"] 				<- level3_last_hrsd_sum
	stardOutputs[stardOutputs$src_subject_id == currentID, "level3_last_hrsd_day"] 				<- level3_last_hrsd_day
	stardOutputs[stardOutputs$src_subject_id == currentID, "level3_remitted"] 				<- level3_remitted
	stardOutputs[stardOutputs$src_subject_id == currentID, "level3_tx"] 					<- level3_tx

	stardOutputs[stardOutputs$src_subject_id == currentID, "level3_hrsdchange"] 				<- level3_hrsdchange
	stardOutputs[stardOutputs$src_subject_id == currentID, "level3_responded"] 				<- level3_responded

	### Now do Level 4 HRSD ###

	if(nrow(currentSubjectHRSD_Level4) == 0){		# If they do not have Level 4, specify everything as NA
		level4_hasHRSDData	<- 0
		level4_last_hrsd_sum	<-  NA
		level4_last_hrsd_day	<-  NA
		level4_remitted 		<-  NA
		level4_hrsdchange		<-  NA
		level4_responded		<-  NA

	}else {			#Otherwise calculate Level 4 data

		level4_hasHRSDData	<- 1

		#Find the latest HRSD day
		level4_last_hrsd_day	<- unique(max(currentSubjectHRSD_Level4$days_baseline))

		# Pull out row matching last day, make sure it is only unique. If multple rows then pull out the first.
		level4_last_hrsd_rows	<- unique(currentSubjectHRSD_Level4[currentSubjectHRSD_Level4$days_baseline == level4_last_hrsd_day& !is.na(currentSubjectHRSD_Level4$days_baseline ) ,])[1,]

		# Pull out last Level 4 hrsd sum
		level4_last_hrsd_sum <- level4_last_hrsd_rows$ hrsdtot_sum_NAasNA

		##Calculate Level 4 remitted: meetInclusion = 1, did not remit in level 1, and Level 4 last HRSD <= 7
		level4_remitted <- ifelse(meetInclusion == 1 & level4_last_hrsd_sum <= 7, 1, 0)

		# Calculate HRSD change
		level4_hrsdchange	<- level4_last_hrsd_sum - level3_last_hrsd_sum

		# Calculate response, defined as 50% improvement from last to first
		level4_responded	<- ifelse((-level4_hrsdchange / level3_last_hrsd_sum) >= 0.50, 1, 0)

	}

	# Pull out level4 treatment
	level4_tx <- ivraP01Raw[ivraP01Raw$src_subject_id == currentID & ivraP01Raw$level == "Level 4" & ivraP01Raw$txassign != "", "txassign"]

	# Convert Level 4 treatment to NA for people who did not get assigned
	level4_tx <- ifelse(length(level4_tx) != 0, level4_tx, NA)

	## Now saving it to output
	stardOutputs[stardOutputs$src_subject_id == currentID, "level4_hasHRSDData"] 				<- level4_hasHRSDData
	stardOutputs[stardOutputs$src_subject_id == currentID, "level4_last_hrsd_sum"] 			<- level4_last_hrsd_sum
	stardOutputs[stardOutputs$src_subject_id == currentID, "level4_last_hrsd_day"] 			<- level4_last_hrsd_day
	stardOutputs[stardOutputs$src_subject_id == currentID, "level4_remitted"] 				<- level4_remitted
	stardOutputs[stardOutputs$src_subject_id == currentID, "level4_tx"] 					<- level4_tx

	stardOutputs[stardOutputs$src_subject_id == currentID, "level4_hrsdchange"] 				<- level4_hrsdchange
	stardOutputs[stardOutputs$src_subject_id == currentID, "level4_responded"] 					<- level4_responded

	########	Adding QIDS Data		########

	#Pull out qids rows for coresponding subject, but only self report
	currentSubjectQids	<- qidsP01Raw[qidsP01Raw$version_form == "Self Rating" & qidsP01Raw$src_subject_id == currentID & !is.na(qidsP01Raw$qstot),]

		if(level1_ivra_entered ==1 ){				##Test if they have entered level 1 according to IVRA
			level1_last_qids_day 	<- max(currentSubjectQids[currentSubjectQids$level == "Level 1", "days_baseline"])
			level1_last_qids_qstot	<- currentSubjectQids[currentSubjectQids$level == "Level 1" & currentSubjectQids$days_baseline == level1_last_qids_day ,"qstot"][1]
			stardOutputs[stardOutputs$src_subject_id == currentID,"level1_last_qids_day"] 	<- level1_last_qids_day
			stardOutputs[stardOutputs$src_subject_id == currentID,"level1_last_qids_qstot"] <- level1_last_qids_qstot

			#Pull date of firstqids
			level1_first_qids_day	<- unique(min(currentSubjectQids[currentSubjectQids$level == "Level 1", "days_baseline"]))
			level1_first_qids_qstot	<- currentSubjectQids[currentSubjectQids$level == "Level 1" & currentSubjectQids$days_baseline == level1_first_qids_day ,"qstot"][1]

			#If they are missing a last HRSD, impute it using the QIDS
			level1_last_hrsd_imputedQIDS  <- ifelse(!is.na(level1_last_hrsd_sum), level1_last_hrsd_sum, remapHRSD_QIDS(level1_last_qids_qstot))
			level1_first_hrsd_sum_imputed <- ifelse(!is.na(level1_first_hrsd_sum), level1_first_hrsd_sum, remapHRSD_QIDS(level1_first_qids_qstot))

			level1_remitted_imputed <- ifelse(meetInclusion == 1 & !is.na(level1_last_hrsd_imputedQIDS  ) & level1_last_hrsd_imputedQIDS  <= 7, 1, 	ifelse(is.na(level1_last_hrsd_imputedQIDS  ), NA, 0))

			level1_hrsdchange_imputed	<- level1_last_hrsd_imputedQIDS   - level1_first_hrsd_sum_imputed
			level1_responded_imputed	<- ifelse((-level1_hrsdchange_imputed / level1_first_hrsd_sum_imputed) >= 0.50, 1, 0)

			stardOutputs[stardOutputs$src_subject_id == currentID,"level1_last_hrsd_imputedQIDS"] 		<- level1_last_hrsd_imputedQIDS
			stardOutputs[stardOutputs$src_subject_id == currentID,"level1_remitted_imputed"] 		<- level1_remitted_imputed
			stardOutputs[stardOutputs$src_subject_id == currentID,"level1_hrsdchange_imputed"] 		<- level1_hrsdchange_imputed
			stardOutputs[stardOutputs$src_subject_id == currentID,"level1_responded_imputed"] 		<- level1_responded_imputed

		}else {
			stardOutputs[stardOutputs$src_subject_id == currentID,"level1_last_qids_day"] 			<- NA
			stardOutputs[stardOutputs$src_subject_id == currentID,"level1_last_qids_qstot"] 		<- NA
			stardOutputs[stardOutputs$src_subject_id == currentID,"level1_last_hrsd_imputedQIDS"] 		<- NA
			stardOutputs[stardOutputs$src_subject_id == currentID,"level1_remitted_imputed"] 		<- NA

			stardOutputs[stardOutputs$src_subject_id == currentID,"level1_hrsdchange_imputed"] 		<- NA
			stardOutputs[stardOutputs$src_subject_id == currentID,"level1_responded_imputed"] 		<- NA

		}

		if(level2_ivra_entered ==1 ){
			level2_last_qids_day 	<- max(currentSubjectQids[currentSubjectQids$level == "Level 2", "days_baseline"])
			level2_last_qids_qstot	<- currentSubjectQids[currentSubjectQids$level == "Level 2" & currentSubjectQids$days_baseline == level2_last_qids_day ,"qstot"][1]
			stardOutputs[stardOutputs$src_subject_id == currentID,"level2_last_qids_day"] 	<- level2_last_qids_day
			stardOutputs[stardOutputs$src_subject_id == currentID,"level2_last_qids_qstot"] <- level2_last_qids_qstot

			level2_last_hrsd_imputedQIDS  <- ifelse(!is.na(level2_last_hrsd_sum), level2_last_hrsd_sum, remapHRSD_QIDS(level2_last_qids_qstot))
			level2_remitted_imputed <- ifelse(meetInclusion == 1 & level2_last_hrsd_imputedQIDS  <= 7, 1, 	ifelse(is.na(level2_last_hrsd_imputedQIDS  ), NA, 0))

			level2_hrsdchange_imputed	<- level2_last_hrsd_imputedQIDS   - level1_last_hrsd_imputedQIDS
			level2_responded_imputed	<- ifelse((-level2_hrsdchange_imputed / level1_last_hrsd_imputedQIDS   ) >= 0.50, 1, 0)

			stardOutputs[stardOutputs$src_subject_id == currentID,"level2_last_hrsd_imputedQIDS"] 		<- level2_last_hrsd_imputedQIDS
			stardOutputs[stardOutputs$src_subject_id == currentID,"level2_remitted_imputed"] 		<- level2_remitted_imputed

			stardOutputs[stardOutputs$src_subject_id == currentID,"level2_hrsdchange_imputed"] 		<- level2_hrsdchange_imputed
			stardOutputs[stardOutputs$src_subject_id == currentID,"level2_responded_imputed"] 		<- level2_responded_imputed

		}else {
			stardOutputs[stardOutputs$src_subject_id == currentID,"level2_last_qids_day"] 	<- NA
			stardOutputs[stardOutputs$src_subject_id == currentID,"level2_last_qids_qstot"] 	<- NA
			stardOutputs[stardOutputs$src_subject_id == currentID,"level2_last_hrsd_imputedQIDS"] 	<- NA
			stardOutputs[stardOutputs$src_subject_id == currentID,"level2_remitted_imputed"] 		<- NA
			stardOutputs[stardOutputs$src_subject_id == currentID,"level2_hrsdchange_imputed"] 	<- NA
			stardOutputs[stardOutputs$src_subject_id == currentID,"level2_responded_imputed"] 		<- NA

		}
		if(level2A_ivra_entered ==1 ){
			level2A_last_qids_day 	<- max(currentSubjectQids[currentSubjectQids$level == "Level 2 A", "days_baseline"])
			level2A_last_qids_qstot	<- currentSubjectQids[currentSubjectQids$level == "Level 2 A" & currentSubjectQids$days_baseline == level2A_last_qids_day ,"qstot"][1]
			stardOutputs[stardOutputs$src_subject_id == currentID,"level2A_last_qids_day"] 	<- level2A_last_qids_day
			stardOutputs[stardOutputs$src_subject_id == currentID,"level2A_last_qids_qstot"] <- level2A_last_qids_qstot

			level2A_last_hrsd_imputedQIDS  <- ifelse(!is.na(level2A_last_hrsd_sum), level2A_last_hrsd_sum, remapHRSD_QIDS(level2A_last_qids_qstot))
			level2A_remitted_imputed <- ifelse(meetInclusion == 1 & level2A_last_hrsd_imputedQIDS  <= 7, 1, ifelse(is.na(level2A_last_hrsd_imputedQIDS  ), NA, 0))

			level2A_hrsdchange_imputed		<- level2A_last_hrsd_imputedQIDS   - level2_last_hrsd_imputedQIDS
			level2A_responded_imputed		<- ifelse((-level2A_hrsdchange_imputed / level2_last_hrsd_imputedQIDS   ) >= 0.50, 1, 0)

			stardOutputs[stardOutputs$src_subject_id == currentID,"level2A_last_hrsd_imputedQIDS"] 	<- level2A_last_hrsd_imputedQIDS
			stardOutputs[stardOutputs$src_subject_id == currentID,"level2A_remitted_imputed"] 		<- level2A_remitted_imputed
			stardOutputs[stardOutputs$src_subject_id == currentID,"level2A_hrsdchange_imputed"] 		<- level2A_hrsdchange_imputed
			stardOutputs[stardOutputs$src_subject_id == currentID,"level2A_responded_imputed"] 		<- level2A_responded_imputed

		}else {
			stardOutputs[stardOutputs$src_subject_id == currentID,"level2A_last_qids_day"] 	<- NA
			stardOutputs[stardOutputs$src_subject_id == currentID,"level2A_last_qids_qstot"] 	<- NA
			stardOutputs[stardOutputs$src_subject_id == currentID,"level2A_last_hrsd_imputedQIDS"] 	<- NA
			stardOutputs[stardOutputs$src_subject_id == currentID,"level2A_remitted_imputed"] 		<- NA

			stardOutputs[stardOutputs$src_subject_id == currentID,"level2A_hrsdchange_imputed"] 	<- NA
			stardOutputs[stardOutputs$src_subject_id == currentID,"level2A_responded_imputed"] 		<- NA

		}

		if(level3_ivra_entered ==1 ){
			level3_last_qids_day 	<- max(currentSubjectQids[currentSubjectQids$level == "Level 3", "days_baseline"])
			level3_last_qids_qstot	<- currentSubjectQids[currentSubjectQids$level == "Level 3" & currentSubjectQids$days_baseline == level3_last_qids_day ,"qstot"][1]
			stardOutputs[stardOutputs$src_subject_id == currentID,"level3_last_qids_day"] 	<- level3_last_qids_day
			stardOutputs[stardOutputs$src_subject_id == currentID,"level3_last_qids_qstot"] <- level3_last_qids_qstot

			level3_last_hrsd_imputedQIDS  <- ifelse(!is.na(level3_last_hrsd_sum), level3_last_hrsd_sum, remapHRSD_QIDS(level3_last_qids_qstot))
			level3_remitted_imputed <- ifelse(meetInclusion == 1 & level3_last_hrsd_imputedQIDS  <= 7, 1, 	ifelse(is.na(level3_last_hrsd_imputedQIDS  ), NA, 0))

			# Create level 3 entry variable depending on if they entered 2A or just 2
			level3_entry_hrsd_sum_imputed	<- ifelse(level2A_ivra_entered == 1, level2A_last_hrsd_imputedQIDS, level2_last_hrsd_imputedQIDS)

			level3_hrsdchange_imputed		<- level3_last_hrsd_imputedQIDS   - level3_entry_hrsd_sum_imputed
			level3_responded_imputed		<- ifelse((-level3_hrsdchange_imputed / level3_entry_hrsd_sum_imputed) >= 0.50, 1, 0)

			stardOutputs[stardOutputs$src_subject_id == currentID,"level3_last_hrsd_imputedQIDS"] 	<- level3_last_hrsd_imputedQIDS
			stardOutputs[stardOutputs$src_subject_id == currentID,"level3_remitted_imputed"] 		<- level3_remitted_imputed
			stardOutputs[stardOutputs$src_subject_id == currentID,"level3_hrsdchange_imputed"] 		<- level3_hrsdchange_imputed
			stardOutputs[stardOutputs$src_subject_id == currentID,"level3_responded_imputed"] 		<- level3_responded_imputed
		}else {
			stardOutputs[stardOutputs$src_subject_id == currentID,"level3_last_qids_day"] 	<- NA
			stardOutputs[stardOutputs$src_subject_id == currentID,"level3_last_qids_qstot"] 	<- NA
			stardOutputs[stardOutputs$src_subject_id == currentID,"level3_last_hrsd_imputedQIDS"] 	<- NA
			stardOutputs[stardOutputs$src_subject_id == currentID,"level3_remitted_imputed"] 		<- NA
			stardOutputs[stardOutputs$src_subject_id == currentID,"level3_hrsdchange_imputed"] 	<- NA
			stardOutputs[stardOutputs$src_subject_id == currentID,"level3_responded_imputed"] 		<- NA
		}
		if(level4_ivra_entered ==1 ){
			level4_last_qids_day 	<- max(currentSubjectQids[currentSubjectQids$level == "Level 4", "days_baseline"])
			level4_last_qids_qstot	<- currentSubjectQids[currentSubjectQids$level == "Level 4" & currentSubjectQids$days_baseline == level4_last_qids_day ,"qstot"][1]
			stardOutputs[stardOutputs$src_subject_id == currentID,"level4_last_qids_day"] 	<- level4_last_qids_day
			stardOutputs[stardOutputs$src_subject_id == currentID,"level4_last_qids_qstot"] <- level4_last_qids_qstot

			level4_last_hrsd_imputedQIDS  <- ifelse(!is.na(level4_last_hrsd_sum), level4_last_hrsd_sum, remapHRSD_QIDS(level4_last_qids_qstot))
			level4_remitted_imputed <- ifelse(meetInclusion == 1 & level4_last_hrsd_imputedQIDS  <= 7, 1, 	ifelse(is.na(level4_last_hrsd_imputedQIDS  ), NA, 0))

			level4_hrsdchange_imputed		<- level4_last_hrsd_imputedQIDS   - level3_last_hrsd_imputedQIDS
			level4_responded_imputed		<- ifelse((-level4_hrsdchange_imputed / level3_last_hrsd_imputedQIDS   ) >= 0.50, 1, 0)

			stardOutputs[stardOutputs$src_subject_id == currentID,"level4_last_hrsd_imputedQIDS"] 		<- level4_last_hrsd_imputedQIDS
			stardOutputs[stardOutputs$src_subject_id == currentID,"level4_remitted_imputed"] 		<- level4_remitted_imputed
			stardOutputs[stardOutputs$src_subject_id == currentID,"level4_hrsdchange_imputed"] 		<- level4_hrsdchange_imputed
			stardOutputs[stardOutputs$src_subject_id == currentID,"level4_responded_imputed"] 		<- level4_responded_imputed

		}else {
			stardOutputs[stardOutputs$src_subject_id == currentID,"level4_last_qids_day"] 	<- NA
			stardOutputs[stardOutputs$src_subject_id == currentID,"level4_last_qids_qstot"] 	<- NA
			stardOutputs[stardOutputs$src_subject_id == currentID,"level4_last_hrsd_imputedQIDS"] 	<- NA
			stardOutputs[stardOutputs$src_subject_id == currentID,"level4_remitted_imputed"] 		<- NA
			stardOutputs[stardOutputs$src_subject_id == currentID,"level4_hrsdchange_imputed"] 	<- NA
			stardOutputs[stardOutputs$src_subject_id == currentID,"level4_responded_imputed"] 		<- NA
		}

		########	Followup Data	########

		# Do they have follow HRSD Data recorded
		fu_hasHRSDData <- ifelse(nrow(currentSubjectHRSD_fu > 0) ,1 ,0)

		if(fu_ivra_entered  == 1){		#Check if they entered followup as recorded on IVRA

			currentSubjectQIDS_fu	<- qidsP01Raw[qidsP01Raw$src_subject_id == currentID& qidsP01Raw$level == "Follow-Up",]

			## Pull out matching followup HRSD for patient
			fu_hrsd_month3		<- currentSubjectHRSD_fu[currentSubjectHRSD_fu$month ==3 & !is.na(currentSubjectHRSD_fu$month),]$hrsdtot_sum_NAasNA
			fu_hrsd_month6		<- currentSubjectHRSD_fu[currentSubjectHRSD_fu$month ==6 & !is.na(currentSubjectHRSD_fu$month),]$hrsdtot_sum_NAasNA
			fu_hrsd_month9		<- currentSubjectHRSD_fu[currentSubjectHRSD_fu$month ==9 & !is.na(currentSubjectHRSD_fu$month),]$hrsdtot_sum_NAasNA
			fu_hrsd_month12		<- currentSubjectHRSD_fu[currentSubjectHRSD_fu$month ==12 & !is.na(currentSubjectHRSD_fu$month),]$hrsdtot_sum_NAasNA

			#Identify observations falling within qids windows

			fu_qids_month3_obs		<- currentSubjectQIDS_fu[currentSubjectQIDS_fu$days_baseline >= qidsWindowMonth3_lo & currentSubjectQIDS_fu$days_baseline  <=  qidsWindowMonth3_hi,]
			fu_qids_month6_obs		<- currentSubjectQIDS_fu[currentSubjectQIDS_fu$days_baseline >= qidsWindowMonth6_lo & currentSubjectQIDS_fu$days_baseline  <=  qidsWindowMonth6_hi,]
			fu_qids_month9_obs		<- currentSubjectQIDS_fu[currentSubjectQIDS_fu$days_baseline >= qidsWindowMonth9_lo & currentSubjectQIDS_fu$days_baseline  <=  qidsWindowMonth9_hi,]
			fu_qids_month12_obs		<- currentSubjectQIDS_fu[currentSubjectQIDS_fu$days_baseline >= qidsWindowMonth12_lo & currentSubjectQIDS_fu$days_baseline  <=  qidsWindowMonth12_hi,]

			fu_qids_month3_minDiff	<- min(abs(fu_qids_month3_obs$days_baseline - qidsWindowMonth3_mid))
			fu_qids_month6_minDiff	<- min(abs(fu_qids_month6_obs$days_baseline - qidsWindowMonth6_mid))
			fu_qids_month9_minDiff	<- min(abs(fu_qids_month9_obs$days_baseline - qidsWindowMonth9_mid))
			fu_qids_month12_minDiff	<- min(abs(fu_qids_month12_obs$days_baseline - qidsWindowMonth12_mid))

			fu_qids_month3_index <- which(abs(fu_qids_month3_obs$days_baseline - qidsWindowMonth3_mid) == fu_qids_month3_minDiff)
			fu_qids_month6_index <- which(abs(fu_qids_month6_obs$days_baseline - qidsWindowMonth6_mid) == fu_qids_month6_minDiff)
			fu_qids_month9_index <- which(abs(fu_qids_month9_obs$days_baseline - qidsWindowMonth9_mid) == fu_qids_month9_minDiff)
			fu_qids_month12_index <- which(abs(fu_qids_month12_obs$days_baseline - qidsWindowMonth12_mid) == fu_qids_month12_minDiff)

			fu_qids_month3		<-	fu_qids_month3_obs[fu_qids_month3_index ,]$SR_TOTAL
			fu_qids_month3_date	<-	fu_qids_month3_obs[fu_qids_month3_index ,]$days_baseline
			fu_qids_month6		<-	fu_qids_month6_obs[fu_qids_month6_index ,]$SR_TOTAL
			fu_qids_month6_date	<-	fu_qids_month6_obs[fu_qids_month6_index ,]$days_baseline
			fu_qids_month9		<-	fu_qids_month9_obs[fu_qids_month9_index ,]$SR_TOTAL
			fu_qids_month9_date	<-	fu_qids_month9_obs[fu_qids_month9_index ,]$days_baseline
			fu_qids_month12		<-	fu_qids_month12_obs[fu_qids_month12_index ,]$SR_TOTAL
			fu_qids_month12_date	<-	fu_qids_month12_obs[fu_qids_month12_index ,]$days_baseline

			# Removing null observations to NA	#

			fu_hrsd_month3	<- ifelse(is.null(fu_hrsd_month3), NA, fu_hrsd_month3)
			fu_hrsd_month6 	<- ifelse(is.null(fu_hrsd_month6), NA, fu_hrsd_month6)
			fu_hrsd_month9	<- ifelse(is.null(fu_hrsd_month9), NA, fu_hrsd_month9)
			fu_hrsd_month12	<- ifelse(is.null(fu_hrsd_month12), NA, fu_hrsd_month12)

			fu_qids_month3	<- ifelse(is.null(fu_qids_month3), NA, fu_qids_month3)
			fu_qids_month6	<- ifelse(is.null(fu_qids_month6), NA, fu_qids_month6)
			fu_qids_month9	<- ifelse(is.null(fu_qids_month9), NA, fu_qids_month9)
			fu_qids_month12	<- ifelse(is.null(fu_qids_month12), NA, fu_qids_month12)

			fu_qids_month3_date	<- ifelse(is.null(fu_qids_month3_date), NA, fu_qids_month3_date)
			fu_qids_month6_date	<- ifelse(is.null(fu_qids_month6_date), NA, fu_qids_month6_date)
			fu_qids_month9_date	<- ifelse(is.null(fu_qids_month9_date), NA, fu_qids_month9_date)
			fu_qids_month12_date	<- ifelse(is.null(fu_qids_month12_date), NA, fu_qids_month12_date)

			# Remapping qids to HRSD where missing HRSD
			fu_hrsdImputed_month3 	<- ifelse(!is.na(fu_hrsd_month3 ), fu_hrsd_month3 , remapHRSD_QIDS(fu_qids_month3))
			fu_hrsdImputed_month6 	<- ifelse(!is.na(fu_hrsd_month6 ), fu_hrsd_month6 , remapHRSD_QIDS(fu_qids_month6))
			fu_hrsdImputed_month9 	<- ifelse(!is.na(fu_hrsd_month9 ), fu_hrsd_month9 , remapHRSD_QIDS(fu_qids_month9))
			fu_hrsdImputed_month12 	<- ifelse(!is.na(fu_hrsd_month12 ), fu_hrsd_month12 , remapHRSD_QIDS(fu_qids_month12))

			# Followup Relapse is defined as any imputed HRSD as >= 14
			fu_relapse <- ifelse( 	(fu_hrsdImputed_month3 >= 14 & !is.na(fu_hrsdImputed_month3)) |
							(fu_hrsdImputed_month6 >= 14 & !is.na(fu_hrsdImputed_month6)) |
							(fu_hrsdImputed_month9 >= 14 & !is.na(fu_hrsdImputed_month9)) |
							(fu_hrsdImputed_month12 >= 14 & !is.na(fu_hrsdImputed_month12)) , 1, 0)

			# Followup Sustained remission FULL is defined as all 4 imputed HRSD <= 7
			fu_susRemission_full <- ifelse( (fu_hrsdImputed_month3 <= 7 &  fu_hrsdImputed_month6 <= 7  & fu_hrsdImputed_month9 <= 7 & fu_hrsdImputed_month12 <= 7) &
								  (!is.na(fu_hrsdImputed_month3) & !is.na(fu_hrsdImputed_month6) & !is.na(fu_hrsdImputed_month9) & !is.na(fu_hrsdImputed_month12)), 1, 0)

			# Followup Sustained remission FULL is defined as all 4 imputed HRSD <= 7 OR MISSING (i.e., no HRSD above > 7)
			fu_susRemission_weak  <- ifelse( (fu_hrsdImputed_month3 <= 7  | is.na(fu_hrsdImputed_month3)) &
								   (fu_hrsdImputed_month6 <= 7  | is.na(fu_hrsdImputed_month6)) &
								   (fu_hrsdImputed_month9 <= 7  | is.na(fu_hrsdImputed_month9)) &
								   (fu_hrsdImputed_month12 <= 7  | is.na(fu_hrsdImputed_month12)) &
								   (!is.na(fu_hrsdImputed_month3) | !is.na(fu_hrsdImputed_month6) | !is.na(fu_hrsdImputed_month9) | !is.na(fu_hrsdImputed_month12)), 1, 0)

			#Check if they entered remittedi in the maxlevelIVRA before remission

			fu_remissionbeforeFU		<- NA
			fu_remissionbeforeFU_imputed	<- NA

			switch(maxlevel_ivra,
					"Level 1" = {fu_remissionbeforeFU		<- level1_remitted},
					"Level 2" = {fu_remissionbeforeFU		<- level2_remitted},
					"Level 2A" = {fu_remissionbeforeFU		<- level2A_remitted},
					"Level 3" = {fu_remissionbeforeFU		<- level3_remitted},
					"Level 4" = {fu_remissionbeforeFU		<- level4_remitted}
				)
			switch(maxlevel_ivra,
					"Level 1" = {fu_remissionbeforeFU_imputed		<- level1_remitted_imputed},
					"Level 2" = {fu_remissionbeforeFU_imputed		<- level2_remitted_imputed},
					"Level 2A" = {fu_remissionbeforeFU_imputed		<- level2A_remitted_imputed},
					"Level 3" = {fu_remissionbeforeFU_imputed		<- level3_remitted_imputed},
					"Level 4" = {fu_remissionbeforeFU_imputed		<- level4_remitted_imputed}
				)

		}else if (fu_ivra_entered == 0){

			fu_hrsd_month3 <- NA
			fu_hrsd_month6 <- NA
			fu_hrsd_month9 <- NA
			fu_hrsd_month12 <- NA
			fu_qids_month3 <- NA
			fu_qids_month6 <- NA
			fu_qids_month9 <- NA
			fu_qids_month12 <- NA
			fu_qids_month3_date <- NA
			fu_qids_month6_date <- NA
			fu_qids_month9_date <- NA
			fu_qids_month12_date <- NA

			fu_hrsdImputed_month3 <- NA
			fu_hrsdImputed_month6 <- NA
			fu_hrsdImputed_month9 <- NA
			fu_hrsdImputed_month12 <- NA

			fu_relapse  <- NA
			fu_susRemission_full <- NA
			fu_susRemission_weak <- NA
			fu_remissionbeforeFU		<- NA
			fu_remissionbeforeFU_imputed	<- NA

		}

		# Calculate if someone has no followup observations

		# Saving followup outputs

		stardOutputs[stardOutputs$src_subject_id == currentID, "fu_hasHRSDData"] <- fu_hasHRSDData

		stardOutputs[stardOutputs$src_subject_id == currentID, "fu_hrsd_month3"] <- fu_hrsd_month3
		stardOutputs[stardOutputs$src_subject_id == currentID, "fu_hrsd_month6"] <- fu_hrsd_month6
		stardOutputs[stardOutputs$src_subject_id == currentID, "fu_hrsd_month9"] <- fu_hrsd_month9
		stardOutputs[stardOutputs$src_subject_id == currentID, "fu_hrsd_month12"] <- fu_hrsd_month12

		stardOutputs[stardOutputs$src_subject_id == currentID, "fu_qids_month3"] <- fu_qids_month3
		stardOutputs[stardOutputs$src_subject_id == currentID, "fu_qids_month6"] <- fu_qids_month6
		stardOutputs[stardOutputs$src_subject_id == currentID, "fu_qids_month9"] <- fu_qids_month9
		stardOutputs[stardOutputs$src_subject_id == currentID, "fu_qids_month12"] <- fu_qids_month12
		stardOutputs[stardOutputs$src_subject_id == currentID, "fu_qids_month3_date"] <- fu_qids_month3_date
		stardOutputs[stardOutputs$src_subject_id == currentID, "fu_qids_month6_date"] <- fu_qids_month6_date
		stardOutputs[stardOutputs$src_subject_id == currentID, "fu_qids_month9_date"] <- fu_qids_month9_date
		stardOutputs[stardOutputs$src_subject_id == currentID, "fu_qids_month12_date"] <- fu_qids_month12_date

		stardOutputs[stardOutputs$src_subject_id == currentID, "fu_hrsdImputed_month3"] <- fu_hrsdImputed_month3
		stardOutputs[stardOutputs$src_subject_id == currentID, "fu_hrsdImputed_month6"] <- fu_hrsdImputed_month6
		stardOutputs[stardOutputs$src_subject_id == currentID, "fu_hrsdImputed_month9"] <- fu_hrsdImputed_month9
		stardOutputs[stardOutputs$src_subject_id == currentID, "fu_hrsdImputed_month12"] <- fu_hrsdImputed_month12

		stardOutputs[stardOutputs$src_subject_id == currentID, "fu_remissionbeforeFU"] 			<- fu_remissionbeforeFU
		stardOutputs[stardOutputs$src_subject_id == currentID, "fu_remissionbeforeFU_imputed"] 	<- fu_remissionbeforeFU_imputed

		stardOutputs[stardOutputs$src_subject_id == currentID, "fu_relapse"] 			<- fu_relapse
		stardOutputs[stardOutputs$src_subject_id == currentID, "fu_susRemission_full"]	<- fu_susRemission_full
		stardOutputs[stardOutputs$src_subject_id == currentID, "fu_susRemission_weak"] 	<- fu_susRemission_weak

	print(i)	# Keep track of progress

}

###########################################################################################
######################			Cleaning up variables		######################
###########################################################################################

##### Which participants entered each level 	####

level1_entered3110 <- stardOutputs[stardOutputs$meetInclusion == 1 & stardOutputs$level1_ivra_entered == 1, "src_subject_id"]
level2_entered1134 <- stardOutputs[stardOutputs$meetInclusion == 1 & (is.na(stardOutputs$level1_remitted ) | stardOutputs$level1_remitted != 1) & stardOutputs$level2_ivra_entered == 1, "src_subject_id"]
level2A_entered28 <- stardOutputs[stardOutputs$meetInclusion == 1 & (is.na(stardOutputs$level1_remitted ) | stardOutputs$level1_remitted != 1) & (is.na(stardOutputs$level2_remitted ) | stardOutputs$level2_remitted != 1) & stardOutputs$level2A_ivra_entered == 1, "src_subject_id"]
level3_entered313 <- stardOutputs[stardOutputs$meetInclusion == 1 & (is.na(stardOutputs$level1_remitted ) | stardOutputs$level1_remitted != 1) & (is.na(stardOutputs$level2_remitted ) | stardOutputs$level2_remitted != 1) & (is.na(stardOutputs$level2A_remitted ) | stardOutputs$level2A_remitted != 1)  & stardOutputs$level3_ivra_entered == 1, "src_subject_id"]
level4_entered94 <- stardOutputs[stardOutputs$meetInclusion == 1 & (is.na(stardOutputs$level1_remitted ) | stardOutputs$level1_remitted != 1) & (is.na(stardOutputs$level2_remitted ) | stardOutputs$level2_remitted != 1) & (is.na(stardOutputs$level2A_remitted ) | stardOutputs$level2A_remitted != 1)  & (is.na(stardOutputs$level3_remitted ) | stardOutputs$level3_remitted != 1)  & stardOutputs$level4_ivra_entered == 1, "src_subject_id"]

############ Create function to calculate mean, 95% CI's and SD's	###############

summarizeOutputs	<- function(x){
	meanX <- mean(x, na.rm = T)
	nX	<- sum(!is.na(x))
	sdX	<- sd(x, na.rm = T)
	seX	<- sdX/sqrt(nX)
	upper95X	<- meanX + 1.96*seX
	lower95X	<- meanX  - 1.96*seX

	summaryOutput <- c(mean = meanX,
						sd = sdX,
						upper95 = upper95X,
						lower95 = lower95X)
	return(summaryOutput)

}

#################################		Calculations for followup 	###################

######	 Create variable for people who have no IVRA followup assessments	##########

# This is how many people entered followup but had NO measurements
stardOutputs$fu_noObservations <- with(stardOutputs, ifelse(is.na(fu_hrsd_month3) & is.na(fu_hrsd_month6) & is.na(fu_hrsd_month9) & is.na(fu_hrsd_month12)
							& is.na(fu_qids_month3) & is.na(fu_qids_month6) & is.na(fu_qids_month9) & is.na(fu_qids_month12), 1,0))

# This is how many people entered followup and had at least ONE measurement
stardOutputs$fu_hasAtLeast1Observations <- with(stardOutputs, ifelse(!is.na(fu_hrsd_month3) | !is.na(fu_hrsd_month6) | !is.na(fu_hrsd_month9) | !is.na(fu_hrsd_month12)
							| !is.na(fu_qids_month3) | !is.na(fu_qids_month6) | !is.na(fu_qids_month9) | !is.na(fu_qids_month12), 1,0))

table(stardOutputs[stardOutputs$meetInclusion == 1 & stardOutputs$fu_ivra_entered == 1,]$fu_noObservations )
table(stardOutputs[stardOutputs$meetInclusion == 1 & stardOutputs$fu_ivra_entered == 1,]$fu_hasAtLeast1Observations )

##### Determining who entered followup before remission		####

# People who entered followup with remission, and then later met relpase

stardOutputs$fu_remit_then_relapse 			<- ifelse(stardOutputs$fu_remissionbeforeFU== 1 & 	stardOutputs$fu_relapse ==1, 1,0)
stardOutputs$fu_NOremit_then_relapse 			<- ifelse(stardOutputs$fu_remissionbeforeFU== 0 & 	stardOutputs$fu_relapse ==1, 1,0)
stardOutputs$fu_remitImputed_then_relapse 		<- ifelse(stardOutputs$fu_remissionbeforeFU_imputed == 1 & 	stardOutputs$fu_relapse ==1, 1,0)
stardOutputs$fu_NOremitImputed_then_relapse 		<- ifelse(stardOutputs$fu_remissionbeforeFU_imputed == 0 & 	stardOutputs$fu_relapse ==1, 1,0)

stardOutputs$fu_remit_then_susRemission_full 			<- ifelse(stardOutputs$fu_remissionbeforeFU == 1 & 	stardOutputs$fu_susRemission_full ==1, 1,0)
stardOutputs$fu_NOremit_then_susRemission_full 		<- ifelse(stardOutputs$fu_remissionbeforeFU == 0 & 	stardOutputs$fu_susRemission_full ==1, 1,0)
stardOutputs$fu_remitImputed_then_susRemission_full 		<- ifelse(stardOutputs$fu_remissionbeforeFU_imputed == 1 & 	stardOutputs$fu_susRemission_full ==1, 1,0)
stardOutputs$fu_NOremitImputed_then_susRemission_full 	<- ifelse(stardOutputs$fu_remissionbeforeFU_imputed == 0 & 	stardOutputs$fu_susRemission_full ==1, 1,0)

stardOutputs$fu_remit_then_susRemission_weak 			<- ifelse(stardOutputs$fu_remissionbeforeFU == 1 & 	stardOutputs$fu_susRemission_weak ==1, 1,0)
stardOutputs$fu_NOremit_then_susRemission_weak 		<- ifelse(stardOutputs$fu_remissionbeforeFU == 0 & 	stardOutputs$fu_susRemission_weak ==1, 1,0)
stardOutputs$fu_remitImputed_then_susRemission_weak 		<- ifelse(stardOutputs$fu_remissionbeforeFU_imputed == 1 & 	stardOutputs$fu_susRemission_weak ==1, 1,0)
stardOutputs$fu_NOremitImputed_then_susRemission_weak 	<- ifelse(stardOutputs$fu_remissionbeforeFU_imputed == 0 & 	stardOutputs$fu_susRemission_weak ==1, 1,0)

##########################			Statistical Tests	 Comparisons	#############

########	ANOVA To compare mean HRSD Change among Level 2 treatments

level2SwitchPatients	<- stardOutputs[stardOutputs$src_subject_id %in% level2_entered1134 & stardOutputs$level2_tx %in% c("SER", "VEN", "BUP"),]

level2AugmentationData	<- stardOutputs[stardOutputs$src_subject_id %in% level2_entered1134 & stardOutputs$level2_tx %in% c("CIT+BUP", "CIT+BUS", "CIT+CT"),]

summary(aov(lm(level2_hrsdchange ~ level2_tx, data = level2SwitchPatients)))
summary(aov(lm(level2_hrsdchange_imputed ~ level2_tx, data = level2SwitchPatients)))

summary(aov(lm(level2_hrsdchange ~ level2_tx, data = level2AugmentationData)))
summary(aov(lm(level2_hrsdchange_imputed ~ level2_tx, data = level2AugmentationData)))

#####################################################################################################
######################			OUTPUTS					######################
#####################################################################################################

## The max level accoring to each person on IVRA
table(stardOutputs$maxlevel_ivra)
table(stardOutputs[stardOutputs$meetInclusion == 1,]$maxlevel_ivra)

###		Calculating how many meet HRSD criteria at DaysBaseline <=5			####

#How many HRSD ≤ 7
table(stardOutputs$level1_first_hrsd_sum	<= 7 , useNA = "always")

#How many 7 < HRSD < 14
table(stardOutputs$level1_first_hrsd_sum<  14 & stardOutputs$level1_first_hrsd_sum> 7 , useNA = "always")

## How many have missing hdtot_sum_NArmT at enrollment
table(is.na(stardOutputs$level1_first_hrsd_sum), useNA = "always")

#####	How many meet inclusion 	###

table(stardOutputs$meetInclusion, useNA = "always")

## How many people entered which level
table(stardOutputs[stardOutputs$meetInclusion == 1,]$level1_ivra_entered, useNA = "always")
table(stardOutputs[stardOutputs$meetInclusion == 1,]$level2_ivra_entered, useNA = "always")
table(stardOutputs[stardOutputs$meetInclusion == 1,]$level2A_ivra_entered, useNA = "always")
table(stardOutputs[stardOutputs$meetInclusion == 1,]$level3_ivra_entered, useNA = "always")
table(stardOutputs[stardOutputs$meetInclusion == 1,]$level4_ivra_entered, useNA = "always")

## How many people entered each level according to IVRA, and met inclusion criteria

nrow(stardOutputs[stardOutputs$meetInclusion == 1 & stardOutputs$level1_ivra_entered == 1,])
nrow(stardOutputs[stardOutputs$meetInclusion == 1 & stardOutputs$level2_ivra_entered == 1,])
nrow(stardOutputs[stardOutputs$meetInclusion == 1 & stardOutputs$level2A_ivra_entered == 1,])
nrow(stardOutputs[stardOutputs$meetInclusion == 1 & stardOutputs$level3_ivra_entered == 1,])
nrow(stardOutputs[stardOutputs$meetInclusion == 1 & stardOutputs$level4_ivra_entered == 1,])

## How many people entered each level according to IVRA, and did not meet remission for previous level , and met inclusion criteria

nrow(stardOutputs[stardOutputs$meetInclusion == 1 & stardOutputs$level1_ivra_entered == 1,])
nrow(stardOutputs[stardOutputs$meetInclusion == 1 & (is.na(stardOutputs$level1_remitted ) | stardOutputs$level1_remitted != 1) & stardOutputs$level2_ivra_entered == 1,])
nrow(stardOutputs[stardOutputs$meetInclusion == 1 & (is.na(stardOutputs$level1_remitted ) | stardOutputs$level1_remitted != 1) & (is.na(stardOutputs$level2_remitted ) | stardOutputs$level2_remitted != 1) & stardOutputs$level2A_ivra_entered == 1,])
nrow(stardOutputs[stardOutputs$meetInclusion == 1 & (is.na(stardOutputs$level1_remitted ) | stardOutputs$level1_remitted != 1) & (is.na(stardOutputs$level2_remitted ) | stardOutputs$level2_remitted != 1) & (is.na(stardOutputs$level2A_remitted ) | stardOutputs$level2A_remitted != 1)  & stardOutputs$level3_ivra_entered == 1,])
nrow(stardOutputs[stardOutputs$meetInclusion == 1 & (is.na(stardOutputs$level1_remitted ) | stardOutputs$level1_remitted != 1) & (is.na(stardOutputs$level2_remitted ) | stardOutputs$level2_remitted != 1) & (is.na(stardOutputs$level2A_remitted ) | stardOutputs$level2A_remitted != 1)  & (is.na(stardOutputs$level3_remitted ) | stardOutputs$level3_remitted != 1)  & stardOutputs$level4_ivra_entered == 1,])

## These are how many people entered each level by treatment

with(stardOutputs[stardOutputs$meetInclusion == 1,] , table(level1_tx, useNA = "always"))
with(stardOutputs[stardOutputs$meetInclusion == 1 & (is.na(stardOutputs$level1_remitted ) | stardOutputs$level1_remitted != 1),] , table(level2_tx, useNA = "always"))
with(stardOutputs[stardOutputs$meetInclusion == 1 & (is.na(stardOutputs$level1_remitted ) | stardOutputs$level1_remitted != 1) & (is.na(stardOutputs$level2_remitted ) | stardOutputs$level2_remitted != 1) & stardOutputs$level2A_ivra_entered == 1,], table(level2A_tx, useNA = "always"))
with(stardOutputs[stardOutputs$meetInclusion == 1 & (is.na(stardOutputs$level1_remitted ) | stardOutputs$level1_remitted != 1) & (is.na(stardOutputs$level2_remitted ) | stardOutputs$level2_remitted != 1) & (is.na(stardOutputs$level2A_remitted ) | stardOutputs$level2A_remitted != 1)  & stardOutputs$level3_ivra_entered == 1,], table(level3_tx, useNA = "always"))
with(stardOutputs[stardOutputs$meetInclusion == 1 & (is.na(stardOutputs$level1_remitted ) | stardOutputs$level1_remitted != 1) & (is.na(stardOutputs$level2_remitted ) | stardOutputs$level2_remitted != 1) & (is.na(stardOutputs$level2A_remitted ) | stardOutputs$level2A_remitted != 1)  & (is.na(stardOutputs$level3_remitted ) | stardOutputs$level3_remitted != 1)  & stardOutputs$level4_ivra_entered == 1,], table(level4_tx, useNA = "always"))

## Now find remissions for each level by treatments ##

with(stardOutputs[stardOutputs$meetInclusion == 1 & stardOutputs$level1_ivra_entered == 1,], table(level1_remitted, level1_tx, useNA = "always"))
with(stardOutputs[stardOutputs$meetInclusion == 1 & (is.na(stardOutputs$level1_remitted ) | stardOutputs$level1_remitted != 1) & stardOutputs$level2_ivra_entered == 1,], table(level2_remitted, level2_tx, useNA = "always"))
with(stardOutputs[stardOutputs$meetInclusion == 1 & (is.na(stardOutputs$level1_remitted ) | stardOutputs$level1_remitted != 1) & (is.na(stardOutputs$level2_remitted ) | stardOutputs$level2_remitted != 1) & stardOutputs$level2A_ivra_entered == 1,], table(level2A_remitted, level2A_tx, useNA = "always"))
with(stardOutputs[stardOutputs$meetInclusion == 1  & (is.na(stardOutputs$level1_remitted ) | stardOutputs$level1_remitted != 1) & (is.na(stardOutputs$level2_remitted ) | stardOutputs$level2_remitted != 1) & (is.na(stardOutputs$level2A_remitted ) | stardOutputs$level2A_remitted != 1)  & stardOutputs$level3_ivra_entered == 1,], table(level3_remitted, level3_tx, useNA = "always"))
with(stardOutputs[stardOutputs$meetInclusion == 1 & (is.na(stardOutputs$level1_remitted ) | stardOutputs$level1_remitted != 1) & (is.na(stardOutputs$level2_remitted ) | stardOutputs$level2_remitted != 1) & (is.na(stardOutputs$level2A_remitted ) | stardOutputs$level2A_remitted != 1)  & (is.na(stardOutputs$level3_remitted ) | stardOutputs$level3_remitted != 1)  & stardOutputs$level4_ivra_entered == 1,], table(level4_remitted, level4_tx, useNA = "always"))

## Remission rates with imputed QIDS

with(stardOutputs[stardOutputs$meetInclusion == 1 & stardOutputs$level1_ivra_entered == 1,], table(level1_remitted_imputed , level1_tx, useNA = "always"))
with(stardOutputs[stardOutputs$meetInclusion == 1 & (is.na(stardOutputs$level1_remitted ) | stardOutputs$level1_remitted != 1) & stardOutputs$level2_ivra_entered == 1,], table(level2_remitted_imputed, level2_tx, useNA = "always"))
with(stardOutputs[stardOutputs$meetInclusion == 1 & (is.na(stardOutputs$level1_remitted ) | stardOutputs$level1_remitted != 1) & (is.na(stardOutputs$level2_remitted ) | stardOutputs$level2_remitted != 1) & stardOutputs$level2A_ivra_entered == 1,], table(level2A_remitted_imputed, level2A_tx, useNA = "always"))
with(stardOutputs[stardOutputs$meetInclusion == 1  & (is.na(stardOutputs$level1_remitted ) | stardOutputs$level1_remitted != 1) & (is.na(stardOutputs$level2_remitted ) | stardOutputs$level2_remitted != 1) & (is.na(stardOutputs$level2A_remitted ) | stardOutputs$level2A_remitted != 1)  & stardOutputs$level3_ivra_entered == 1,], table(level3_remitted_imputed, level3_tx, useNA = "always"))
with(stardOutputs[stardOutputs$meetInclusion == 1 & (is.na(stardOutputs$level1_remitted ) | stardOutputs$level1_remitted != 1) & (is.na(stardOutputs$level2_remitted ) | stardOutputs$level2_remitted != 1) & (is.na(stardOutputs$level2A_remitted ) | stardOutputs$level2A_remitted != 1)  & (is.na(stardOutputs$level3_remitted ) | stardOutputs$level3_remitted != 1)  & stardOutputs$level4_ivra_entered == 1,], table(level4_remitted_imputed, level4_tx, useNA = "always"))

## Now find responses for each level by treatments ##

with(stardOutputs[stardOutputs$meetInclusion == 1 & stardOutputs$level1_ivra_entered == 1,], table(level1_responded, level1_tx, useNA = "always"))
with(stardOutputs[stardOutputs$meetInclusion == 1 & (is.na(stardOutputs$level1_remitted ) | stardOutputs$level1_remitted != 1) & stardOutputs$level2_ivra_entered == 1,], table(level2_responded, level2_tx, useNA = "always"))
with(stardOutputs[stardOutputs$meetInclusion == 1 & (is.na(stardOutputs$level1_remitted ) | stardOutputs$level1_remitted != 1) & (is.na(stardOutputs$level2_remitted ) | stardOutputs$level2_remitted != 1) & stardOutputs$level2A_ivra_entered == 1,], table(level2A_responded, level2A_tx, useNA = "always"))
with(stardOutputs[stardOutputs$meetInclusion == 1  & (is.na(stardOutputs$level1_remitted ) | stardOutputs$level1_remitted != 1) & (is.na(stardOutputs$level2_remitted ) | stardOutputs$level2_remitted != 1) & (is.na(stardOutputs$level2A_remitted ) | stardOutputs$level2A_remitted != 1)  & stardOutputs$level3_ivra_entered == 1,], table(level3_responded, level3_tx, useNA = "always"))
with(stardOutputs[stardOutputs$meetInclusion == 1 & (is.na(stardOutputs$level1_remitted ) | stardOutputs$level1_remitted != 1) & (is.na(stardOutputs$level2_remitted ) | stardOutputs$level2_remitted != 1) & (is.na(stardOutputs$level2A_remitted ) | stardOutputs$level2A_remitted != 1)  & (is.na(stardOutputs$level3_remitted ) | stardOutputs$level3_remitted != 1)  & stardOutputs$level4_ivra_entered == 1,], table(level4_responded, level4_tx, useNA = "always"))

## Now find imputed responses for each level by treatments ##

with(stardOutputs[stardOutputs$meetInclusion == 1 & stardOutputs$level1_ivra_entered == 1,], table(level1_responded_imputed, level1_tx, useNA = "always"))
with(stardOutputs[stardOutputs$meetInclusion == 1 & (is.na(stardOutputs$level1_remitted ) | stardOutputs$level1_remitted != 1) & stardOutputs$level2_ivra_entered == 1,], table(level2_responded_imputed, level2_tx, useNA = "always"))
with(stardOutputs[stardOutputs$meetInclusion == 1 & (is.na(stardOutputs$level1_remitted ) | stardOutputs$level1_remitted != 1) & (is.na(stardOutputs$level2_remitted ) | stardOutputs$level2_remitted != 1) & stardOutputs$level2A_ivra_entered == 1,], table(level2A_responded_imputed, level2A_tx, useNA = "always"))
with(stardOutputs[stardOutputs$meetInclusion == 1  & (is.na(stardOutputs$level1_remitted ) | stardOutputs$level1_remitted != 1) & (is.na(stardOutputs$level2_remitted ) | stardOutputs$level2_remitted != 1) & (is.na(stardOutputs$level2A_remitted ) | stardOutputs$level2A_remitted != 1)  & stardOutputs$level3_ivra_entered == 1,], table(level3_responded_imputed, level3_tx, useNA = "always"))
with(stardOutputs[stardOutputs$meetInclusion == 1 & (is.na(stardOutputs$level1_remitted ) | stardOutputs$level1_remitted != 1) & (is.na(stardOutputs$level2_remitted ) | stardOutputs$level2_remitted != 1) & (is.na(stardOutputs$level2A_remitted ) | stardOutputs$level2A_remitted != 1)  & (is.na(stardOutputs$level3_remitted ) | stardOutputs$level3_remitted != 1)  & stardOutputs$level4_ivra_entered == 1,], table(level4_responded_imputed, level4_tx, useNA = "always"))

## Now find mean HRSD change score  ##

with(stardOutputs[stardOutputs$meetInclusion == 1 & stardOutputs$level1_ivra_entered == 1,], aggregate(level1_hrsdchange, list(level1_tx), mean, na.rm = T))
with(stardOutputs[stardOutputs$meetInclusion == 1 & (is.na(stardOutputs$level1_remitted ) | stardOutputs$level1_remitted != 1) & stardOutputs$level2_ivra_entered == 1,], aggregate(level2_hrsdchange, list(level2_tx), mean, na.rm = T))
with(stardOutputs[stardOutputs$meetInclusion == 1 & (is.na(stardOutputs$level1_remitted ) | stardOutputs$level1_remitted != 1) & (is.na(stardOutputs$level2_remitted ) | stardOutputs$level2_remitted != 1) & stardOutputs$level2A_ivra_entered == 1,], aggregate(level2A_hrsdchange, list(level2A_tx), mean, na.rm = T))
with(stardOutputs[stardOutputs$meetInclusion == 1  & (is.na(stardOutputs$level1_remitted ) | stardOutputs$level1_remitted != 1) & (is.na(stardOutputs$level2_remitted ) | stardOutputs$level2_remitted != 1) & (is.na(stardOutputs$level2A_remitted ) | stardOutputs$level2A_remitted != 1)  & stardOutputs$level3_ivra_entered == 1,], aggregate(level3_hrsdchange, list(level3_tx), mean, na.rm = T))
with(stardOutputs[stardOutputs$meetInclusion == 1 & (is.na(stardOutputs$level1_remitted ) | stardOutputs$level1_remitted != 1) & (is.na(stardOutputs$level2_remitted ) | stardOutputs$level2_remitted != 1) & (is.na(stardOutputs$level2A_remitted ) | stardOutputs$level2A_remitted != 1)  & (is.na(stardOutputs$level3_remitted ) | stardOutputs$level3_remitted != 1)  & stardOutputs$level4_ivra_entered == 1,], aggregate(level4_hrsdchange, list(level4_tx), mean, na.rm = T))

## Now find mean HRSD change imputed score  ##

with(stardOutputs[stardOutputs$meetInclusion == 1 & stardOutputs$level1_ivra_entered == 1,], aggregate(level1_hrsdchange_imputed, list(level1_tx), summarizeOutputs))
with(stardOutputs[stardOutputs$meetInclusion == 1 & (is.na(stardOutputs$level1_remitted ) | stardOutputs$level1_remitted != 1) & stardOutputs$level2_ivra_entered == 1,], aggregate(level2_hrsdchange_imputed, list(level2_tx),  summarizeOutputs))
with(stardOutputs[stardOutputs$meetInclusion == 1 & (is.na(stardOutputs$level1_remitted ) | stardOutputs$level1_remitted != 1) & (is.na(stardOutputs$level2_remitted ) | stardOutputs$level2_remitted != 1) & stardOutputs$level2A_ivra_entered == 1,], aggregate(level2A_hrsdchange_imputed, list(level2A_tx),  summarizeOutputs))
with(stardOutputs[stardOutputs$meetInclusion == 1  & (is.na(stardOutputs$level1_remitted ) | stardOutputs$level1_remitted != 1) & (is.na(stardOutputs$level2_remitted ) | stardOutputs$level2_remitted != 1) & (is.na(stardOutputs$level2A_remitted ) | stardOutputs$level2A_remitted != 1)  & stardOutputs$level3_ivra_entered == 1,], aggregate(level3_hrsdchange_imputed, list(level3_tx),  summarizeOutputs))
with(stardOutputs[stardOutputs$meetInclusion == 1 & (is.na(stardOutputs$level1_remitted ) | stardOutputs$level1_remitted != 1) & (is.na(stardOutputs$level2_remitted ) | stardOutputs$level2_remitted != 1) & (is.na(stardOutputs$level2A_remitted ) | stardOutputs$level2A_remitted != 1)  & (is.na(stardOutputs$level3_remitted ) | stardOutputs$level3_remitted != 1)  & stardOutputs$level4_ivra_entered == 1,], aggregate(level4_hrsdchange_imputed, list(level4_tx),  summarizeOutputs))

##################################################################################################################################
############################				FOLLOWUP CALCULATIONS				########################################
##################################################################################################################################

## Who entered according to IVRA each level	##
with(stardOutputs[stardOutputs$meetInclusion == 1,], table(level1_ivra_entered, useNA = "always"))
with(stardOutputs[stardOutputs$meetInclusion == 1 & (stardOutputs$level1_remitted !=  1  | is.na(stardOutputs$level1_remitted)),], table(level2_ivra_entered, useNA = "always"))
with(stardOutputs[stardOutputs$meetInclusion == 1 & (stardOutputs$level1_remitted !=  1  | is.na(stardOutputs$level1_remitted)) & (stardOutputs$level2_remitted !=  1  | is.na(stardOutputs$level2_remitted)),], table(level2A_ivra_entered, useNA = "always"))
with(stardOutputs[stardOutputs$meetInclusion == 1 & (is.na(stardOutputs$level1_remitted ) | stardOutputs$level1_remitted != 1) & (is.na(stardOutputs$level2_remitted ) | stardOutputs$level2_remitted != 1) & (is.na(stardOutputs$level2A_remitted ) | stardOutputs$level2A_remitted != 1),], table(level3_ivra_entered , useNA = "always"))
with(stardOutputs[stardOutputs$meetInclusion == 1 & (is.na(stardOutputs$level1_remitted ) | stardOutputs$level1_remitted != 1) & (is.na(stardOutputs$level2_remitted ) | stardOutputs$level2_remitted != 1) & (is.na(stardOutputs$level2A_remitted ) | stardOutputs$level2A_remitted != 1)  & (is.na(stardOutputs$level3_remitted ) | stardOutputs$level3_remitted != 1),], table(level4_ivra_entered , useNA = "always"))

## Max level IVRA for each 	##
with(stardOutputs, table(maxlevel_ivra, useNA = "always"))
with(stardOutputs[stardOutputs$meetInclusion == 1,], table(maxlevel_ivra, useNA = "always"))

##################

## How many entered followup in each level according to IVRA - This preserves the correct denominator and uses whether they have followup IVRA
with(stardOutputs[stardOutputs$meetInclusion == 1 & !(stardOutputs$src_subject_id %in% c(level2_entered1134 , level2A_entered28, level3_entered313, level4_entered94)),], table(fu_ivra_entered, useNA = "always"))
with(stardOutputs[stardOutputs$src_subject_id %in% level2_entered1134 & !(stardOutputs$src_subject_id %in% c(level2A_entered28, level3_entered313, level4_entered94)),], table(fu_ivra_entered, useNA = "always"))
with(stardOutputs[stardOutputs$src_subject_id %in% level2A_entered28 & !(stardOutputs$src_subject_id %in% c(level3_entered313, level4_entered94)),], table(fu_ivra_entered, useNA = "always"))
with(stardOutputs[stardOutputs$src_subject_id %in% level3_entered313 & !(stardOutputs$src_subject_id %in% c(level4_entered94)),], table(fu_ivra_entered, useNA = "always"))
with(stardOutputs[stardOutputs$src_subject_id %in% level4_entered94,], table(fu_ivra_entered, useNA = "always"))

## How many entered followup in each level according to IVRA - This preserves the correct denominator and uses whether they have followup IVRA
with(stardOutputs[stardOutputs$meetInclusion == 1 & !(stardOutputs$src_subject_id %in% c(level2_entered1134 , level2A_entered28, level3_entered313, level4_entered94)),], table(fu_ivra_entered, useNA = "always"))
with(stardOutputs[stardOutputs$src_subject_id %in% level2_entered1134 & !(stardOutputs$src_subject_id %in% c(level2A_entered28, level3_entered313, level4_entered94)),], table(fu_ivra_entered, useNA = "always"))
with(stardOutputs[stardOutputs$src_subject_id %in% level2A_entered28 & stardOutputs$level1_remitted != 1  & !(stardOutputs$src_subject_id %in% c(level3_entered313, level4_entered94)),], table(fu_ivra_entered, useNA = "always"))
with(stardOutputs[stardOutputs$src_subject_id %in% level3_entered313 & stardOutputs$level1_remitted != 1  & !(stardOutputs$src_subject_id %in% c(level4_entered94)),], table(fu_ivra_entered, useNA = "always"))
with(stardOutputs[stardOutputs$src_subject_id %in% level4_entered94,], table(fu_ivra_entered, useNA = "always"))

## How many have followup HRSD at each level - This preserves the correct deminator and uses whether they have followup HRSD
with(stardOutputs[stardOutputs$meetInclusion == 1 & !(stardOutputs$src_subject_id %in% c(level2_entered1134 , level2A_entered28, level3_entered313, level4_entered94)),], table(fu_hasHRSDData , useNA = "always"))
with(stardOutputs[stardOutputs$src_subject_id %in% level2_entered1134 & !(stardOutputs$src_subject_id %in% c(level2A_entered28, level3_entered313, level4_entered94)),], table(fu_hasHRSDData , useNA = "always"))
with(stardOutputs[stardOutputs$src_subject_id %in% level2A_entered28 & !(stardOutputs$src_subject_id %in% c(level3_entered313, level4_entered94)),], table(fu_hasHRSDData , useNA = "always"))
with(stardOutputs[stardOutputs$src_subject_id %in% level3_entered313 & !(stardOutputs$src_subject_id %in% c(level4_entered94)),], table(fu_hasHRSDData , useNA = "always"))
with(stardOutputs[stardOutputs$src_subject_id %in% level4_entered94,], table(fu_hasHRSDData , useNA = "always"))

## How many hve followup HRSD according to max IVRA -- This may lead to inconsistencies, because individuals could have been incorrectly moved on via IVRA despite meeting remission on HRSD.
with(stardOutputs[stardOutputs$meetInclusion == 1 & stardOutputs$maxlevel_ivra == "Level 1",], table(fu_hasHRSDData, useNA = "always"))
with(stardOutputs[stardOutputs$meetInclusion == 1 & stardOutputs$maxlevel_ivra == "Level 2",], table(fu_hasHRSDData, useNA = "always"))
with(stardOutputs[stardOutputs$meetInclusion == 1 & stardOutputs$maxlevel_ivra == "Level 2A",], table(fu_hasHRSDData, useNA = "always"))
with(stardOutputs[stardOutputs$meetInclusion == 1 & stardOutputs$maxlevel_ivra == "Level 3",], table(fu_hasHRSDData, useNA = "always"))
with(stardOutputs[stardOutputs$meetInclusion == 1 & stardOutputs$maxlevel_ivra == "Level 4",], table(fu_hasHRSDData, useNA = "always"))

###### Calculating sustained remission	########

## How many entered followup in each level according to IVRA - This preserves the correct denominator and uses whether they have followup IVRA
with(stardOutputs[stardOutputs$meetInclusion == 1 & !(stardOutputs$src_subject_id %in% c(level2_entered1134 , level2A_entered28, level3_entered313, level4_entered94)),], table(fu_ivra_entered, level1_tx, useNA = "always"))
with(stardOutputs[stardOutputs$meetInclusion == 1 & !(stardOutputs$src_subject_id %in% c(level2_entered1134 , level2A_entered28, level3_entered313, level4_entered94)),], table(fu_relapse, level1_tx, useNA = "always"))
with(stardOutputs[stardOutputs$meetInclusion == 1 & !(stardOutputs$src_subject_id %in% c(level2_entered1134 , level2A_entered28, level3_entered313, level4_entered94)),], table(fu_susRemission_full, level1_tx, useNA = "always"))
with(stardOutputs[stardOutputs$meetInclusion == 1 & !(stardOutputs$src_subject_id %in% c(level2_entered1134 , level2A_entered28, level3_entered313, level4_entered94)),], table(fu_susRemission_weak, level1_tx, useNA = "always"))

with(stardOutputs[stardOutputs$src_subject_id %in% level2_entered1134 & !(stardOutputs$src_subject_id %in% c(level2A_entered28, level3_entered313, level4_entered94)),], table(fu_ivra_entered, level2_tx, useNA = "always"))
with(stardOutputs[stardOutputs$src_subject_id %in% level2_entered1134 & !(stardOutputs$src_subject_id %in% c(level2A_entered28, level3_entered313, level4_entered94)),], table(fu_relapse, level2_tx, useNA = "always"))
with(stardOutput	s[stardOutputs$src_subject_id %in% level2_entered1134 & !(stardOutputs$src_subject_id %in% c(level2A_entered28, level3_entered313, level4_entered94)),], table(fu_susRemission_full, level2_tx, useNA = "always"))
with(stardOutputs[stardOutputs$src_subject_id %in% level2_entered1134 & !(stardOutputs$src_subject_id %in% c(level2A_entered28, level3_entered313, level4_entered94)),], table(fu_susRemission_weak, level2_tx, useNA = "always"))

with(stardOutputs[stardOutputs$src_subject_id %in% level2A_entered28 & !(stardOutputs$src_subject_id %in% c(level3_entered313, 13, level4_entered94)),], table(fu_ivra_entered, level2A_tx, useNA = "always"))
with(stardOutputs[stardOutputs$src_subject_id %in% level2A_entered28 & !(stardOutputs$src_subject_id %in% c(level3_entered313, 13, level4_entered94)),], table(fu_relapse, level2A_tx, useNA = "always"))
with(stardOutputs[stardOutputs$src_subject_id %in% level2A_entered28 & !(stardOutputs$src_subject_id %in% c(level3_entered313, 13, level4_entered94)),], table(fu_susRemission_full, level2A_tx, useNA = "always"))
with(stardOutputs[stardOutputs$src_subject_id %in% level2A_entered28 & !(stardOutputs$src_subject_id %in% c(level3_entered313, 13, level4_entered94)),], table(fu_susRemission_weak, level2A_tx, useNA = "always"))

with(stardOutputs[stardOutputs$src_subject_id %in% level3_entered313 & !(stardOutputs$src_subject_id %in% c(level4_entered94)),], table(fu_ivra_entered, level3_tx, useNA = "always"))
with(stardOutputs[stardOutputs$src_subject_id %in% level3_entered313 & !(stardOutputs$src_subject_id %in% c(level4_entered94)),], table(fu_relapse, level3_tx, useNA = "always"))
with(stardOutputs[stardOutputs$src_subject_id %in% level3_entered313 & !(stardOutputs$src_subject_id %in% c(level4_entered94)),], table(fu_susRemission_full, level3_tx, useNA = "always"))
with(stardOutputs[stardOutputs$src_subject_id %in% level3_entered313 & !(stardOutputs$src_subject_id %in% c(level4_entered94)),], table(fu_susRemission_weak, level3_tx, useNA = "always"))

with(stardOutputs[stardOutputs$src_subject_id %in% level4_entered94,], table(fu_ivra_entered, level4_tx, useNA = "always"))
with(stardOutputs[stardOutputs$src_subject_id %in% level4_entered94,], table(fu_relapse, level4_tx, useNA = "always"))
with(stardOutputs[stardOutputs$src_subject_id %in% level4_entered94,], table(fu_susRemission_full, level4_tx, useNA = "always"))
with(stardOutputs[stardOutputs$src_subject_id %in% level4_entered94,], table(fu_susRemission_weak, level4_tx, useNA = "always"))

##########################################################################################################################
########################			COMBINING INTO A SINGLE OUTPUT			################################
##########################################################################################################################

## Level 1	##
(outputs_level1_count				<- with(stardOutputs[stardOutputs$meetInclusion == 1,] , table(level1_tx, useNA = "always")))
(outputs_level1_remitted				<- with(stardOutputs[stardOutputs$meetInclusion == 1 & stardOutputs$level1_ivra_entered == 1,], table(level1_remitted, level1_tx, useNA = "always")))
(outputs_level1_remitted_imputed			<- with(stardOutputs[stardOutputs$meetInclusion == 1 & stardOutputs$level1_ivra_entered == 1,], table(level1_remitted_imputed , level1_tx, useNA = "always")))
(outputs_level1_responded				<- with(stardOutputs[stardOutputs$meetInclusion == 1 & stardOutputs$level1_ivra_entered == 1,], table(level1_responded, level1_tx, useNA = "always")))
(outputs_level1_responded_imputed			<- with(stardOutputs[stardOutputs$meetInclusion == 1 & stardOutputs$level1_ivra_entered == 1,], table(level1_responded_imputed , level1_tx, useNA = "always")))
(outputs_level1_fu_ivra_entered			<- with(stardOutputs[stardOutputs$meetInclusion == 1 & !(stardOutputs$src_subject_id %in% c(level2_entered1134 , level2A_entered28, level3_entered313, level4_entered94)),], table(fu_ivra_entered, level1_tx, useNA = "always")))
(outputs_level1_fu_relapse				<- with(stardOutputs[stardOutputs$meetInclusion == 1 & !(stardOutputs$src_subject_id %in% c(level2_entered1134 , level2A_entered28, level3_entered313, level4_entered94)),], table(fu_relapse, level1_tx, useNA = "always")))
(outputs_level1_fu_susRemission_full		<- with(stardOutputs[stardOutputs$meetInclusion == 1 & !(stardOutputs$src_subject_id %in% c(level2_entered1134 , level2A_entered28, level3_entered313, level4_entered94)),], table(fu_susRemission_full, level1_tx, useNA = "always")))
(outputs_level1_fu_susRemission_weak		<- with(stardOutputs[stardOutputs$meetInclusion == 1 & !(stardOutputs$src_subject_id %in% c(level2_entered1134 , level2A_entered28, level3_entered313, level4_entered94)),], table(fu_susRemission_weak, level1_tx, useNA = "always")))
(outputs_level1_fu_hasAtLeast1Observations	<- with(stardOutputs[stardOutputs$meetInclusion == 1 & !(stardOutputs$src_subject_id %in% c(level2_entered1134 , level2A_entered28, level3_entered313, level4_entered94)),], table(fu_hasAtLeast1Observations , level1_tx, useNA = "always")))

(outputs_level1_fu_remissionbeforeFU				<- with(stardOutputs[stardOutputs$meetInclusion == 1 & !(stardOutputs$src_subject_id %in% c(level2_entered1134 , level2A_entered28, level3_entered313, level4_entered94)),], table(fu_remissionbeforeFU, level1_tx, useNA = "always")))
(outputs_level1_fu_remissionbeforeFU_imputed			<- with(stardOutputs[stardOutputs$meetInclusion == 1 & !(stardOutputs$src_subject_id %in% c(level2_entered1134 , level2A_entered28, level3_entered313, level4_entered94)),], table(fu_remissionbeforeFU_imputed, level1_tx, useNA = "always")))
(outputs_level1_fu_remitImputed_then_relapse			<- with(stardOutputs[stardOutputs$meetInclusion == 1 & !(stardOutputs$src_subject_id %in% c(level2_entered1134 , level2A_entered28, level3_entered313, level4_entered94)),], table(fu_remitImputed_then_relapse, level1_tx, useNA = "always")))
(outputs_level1_fu_NOremitImputed_then_relapse 		<- with(stardOutputs[stardOutputs$meetInclusion == 1 & !(stardOutputs$src_subject_id %in% c(level2_entered1134 , level2A_entered28, level3_entered313, level4_entered94)),], table(fu_NOremitImputed_then_relapse , level1_tx, useNA = "always")))
(outputs_level1_fu_remitImputed_then_susRemission_full 	<- with(stardOutputs[stardOutputs$meetInclusion == 1 & !(stardOutputs$src_subject_id %in% c(level2_entered1134 , level2A_entered28, level3_entered313, level4_entered94)),], table(fu_remitImputed_then_susRemission_full , level1_tx, useNA = "always")))
(outputs_level1_fu_NOremitImputed_then_susRemission_full	<- with(stardOutputs[stardOutputs$meetInclusion == 1 & !(stardOutputs$src_subject_id %in% c(level2_entered1134 , level2A_entered28, level3_entered313, level4_entered94)),], table(fu_NOremitImputed_then_susRemission_full, level1_tx, useNA = "always")))
(outputs_level1_fu_remitImputed_then_susRemission_weak	<- with(stardOutputs[stardOutputs$meetInclusion == 1 & !(stardOutputs$src_subject_id %in% c(level2_entered1134 , level2A_entered28, level3_entered313, level4_entered94)),], table(fu_remitImputed_then_susRemission_weak, level1_tx, useNA = "always")))
(outputs_level1_fu_NOremitImputed_then_susRemission_weak	<- with(stardOutputs[stardOutputs$meetInclusion == 1 & !(stardOutputs$src_subject_id %in% c(level2_entered1134 , level2A_entered28, level3_entered313, level4_entered94)),], table(fu_NOremitImputed_then_susRemission_weak, level1_tx, useNA = "always")))

## Level 2	##
(outputs_level2_count					<- with(stardOutputs[stardOutputs$meetInclusion == 1 & (is.na(stardOutputs$level1_remitted ) | stardOutputs$level1_remitted != 1),] , table(level2_tx, useNA = "always")))
(outputs_level2_remitted				<- with(stardOutputs[stardOutputs$meetInclusion == 1 & (is.na(stardOutputs$level1_remitted ) | stardOutputs$level1_remitted != 1) & stardOutputs$level2_ivra_entered == 1,], table(level2_remitted, level2_tx, useNA = "always")))
(outputs_level2_remitted_imputed			<- with(stardOutputs[stardOutputs$meetInclusion == 1 & (is.na(stardOutputs$level1_remitted ) | stardOutputs$level1_remitted != 1) & stardOutputs$level2_ivra_entered == 1,], table(level2_remitted_imputed, level2_tx, useNA = "always")))
(outputs_level2_responded				<- with(stardOutputs[stardOutputs$meetInclusion == 1 & (is.na(stardOutputs$level1_remitted ) | stardOutputs$level1_remitted != 1) & stardOutputs$level2_ivra_entered == 1,], table(level2_responded, level2_tx, useNA = "always")))
(outputs_level2_responded_imputed			<- with(stardOutputs[stardOutputs$meetInclusion == 1 & (is.na(stardOutputs$level1_remitted ) | stardOutputs$level1_remitted != 1) & stardOutputs$level2_ivra_entered == 1,], table(level2_responded_imputed, level2_tx, useNA = "always")))
(outputs_level2_fu_ivra_entered			<- with(stardOutputs[stardOutputs$src_subject_id %in% level2_entered1134 & !(stardOutputs$src_subject_id %in% c(level2A_entered28, level3_entered313, level4_entered94)),], table(fu_ivra_entered, level2_tx, useNA = "always")))
(outputs_level2_fu_relapse				<- with(stardOutputs[stardOutputs$src_subject_id %in% level2_entered1134 & !(stardOutputs$src_subject_id %in% c(level2A_entered28, level3_entered313, level4_entered94)),], table(fu_relapse, level2_tx, useNA = "always")))
(outputs_level2_fu_susRemission_full		<- with(stardOutputs[stardOutputs$src_subject_id %in% level2_entered1134 & !(stardOutputs$src_subject_id %in% c(level2A_entered28, level3_entered313, level4_entered94)),], table(fu_susRemission_full, level2_tx, useNA = "always")))
(outputs_level2_fu_susRemission_weak		<- with(stardOutputs[stardOutputs$src_subject_id %in% level2_entered1134 & !(stardOutputs$src_subject_id %in% c(level2A_entered28, level3_entered313, level4_entered94)),], table(fu_susRemission_weak, level2_tx, useNA = "always")))
(outputs_level2_fu_hasAtLeast1Observations 	<- with(stardOutputs[stardOutputs$src_subject_id %in% level2_entered1134 & !(stardOutputs$src_subject_id %in% c(level2A_entered28, level3_entered313, level4_entered94)),], table(fu_hasAtLeast1Observations , level2_tx, useNA = "always")))

(outputs_level2_fu_remissionbeforeFU				<- with(stardOutputs[stardOutputs$src_subject_id %in% level2_entered1134 & !(stardOutputs$src_subject_id %in% c(level2A_entered28, level3_entered313, level4_entered94)),], table(fu_remissionbeforeFU, level2_tx, useNA = "always")))
(outputs_level2_fu_remissionbeforeFU_imputed			<- with(stardOutputs[stardOutputs$src_subject_id %in% level2_entered1134 & !(stardOutputs$src_subject_id %in% c(level2A_entered28, level3_entered313, level4_entered94)),], table(fu_remissionbeforeFU_imputed, level2_tx, useNA = "always")))
(outputs_level2_fu_remitImputed_then_relapse			<- with(stardOutputs[stardOutputs$src_subject_id %in% level2_entered1134 & !(stardOutputs$src_subject_id %in% c(level2A_entered28, level3_entered313, level4_entered94)),], table(fu_remitImputed_then_relapse, level2_tx, useNA = "always")))
(outputs_level2_fu_NOremitImputed_then_relapse 		<- with(stardOutputs[stardOutputs$src_subject_id %in% level2_entered1134 & !(stardOutputs$src_subject_id %in% c(level2A_entered28, level3_entered313, level4_entered94)),], table(fu_NOremitImputed_then_relapse , level2_tx, useNA = "always")))
(outputs_level2_fu_remitImputed_then_susRemission_full 	<- with(stardOutputs[stardOutputs$src_subject_id %in% level2_entered1134 & !(stardOutputs$src_subject_id %in% c(level2A_entered28, level3_entered313, level4_entered94)),], table(fu_remitImputed_then_susRemission_full , level2_tx, useNA = "always")))
(outputs_level2_fu_NOremitImputed_then_susRemission_full	<- with(stardOutputs[stardOutputs$src_subject_id %in% level2_entered1134 & !(stardOutputs$src_subject_id %in% c(level2A_entered28, level3_entered313, level4_entered94)),], table(fu_NOremitImputed_then_susRemission_full, level2_tx, useNA = "always")))
(outputs_level2_fu_remitImputed_then_susRemission_weak	<- with(stardOutputs[stardOutputs$src_subject_id %in% level2_entered1134 & !(stardOutputs$src_subject_id %in% c(level2A_entered28, level3_entered313, level4_entered94)),], table(fu_remitImputed_then_susRemission_weak, level2_tx, useNA = "always")))
(outputs_level2_fu_NOremitImputed_then_susRemission_weak	<- with(stardOutputs[stardOutputs$src_subject_id %in% level2_entered1134 & !(stardOutputs$src_subject_id %in% c(level2A_entered28, level3_entered313, level4_entered94)),], table(fu_NOremitImputed_then_susRemission_weak, level2_tx, useNA = "always")))

## Level 2A	##
(outputs_level2A_count					<- with(stardOutputs[stardOutputs$meetInclusion == 1 & (is.na(stardOutputs$level1_remitted ) | stardOutputs$level1_remitted != 1) & (is.na(stardOutputs$level2_remitted ) | stardOutputs$level2_remitted != 1) & stardOutputs$level2A_ivra_entered == 1,], table(level2A_tx, useNA = "always")))
(outputs_level2A_remitted				<- with(stardOutputs[stardOutputs$meetInclusion == 1 & (is.na(stardOutputs$level1_remitted ) | stardOutputs$level1_remitted != 1) & (is.na(stardOutputs$level2_remitted ) | stardOutputs$level2_remitted != 1) & stardOutputs$level2A_ivra_entered == 1,], table(level2A_remitted, level2A_tx, useNA = "always")))
(outputs_level2A_remitted_imputed			<- with(stardOutputs[stardOutputs$meetInclusion == 1 & (is.na(stardOutputs$level1_remitted ) | stardOutputs$level1_remitted != 1) & (is.na(stardOutputs$level2_remitted ) | stardOutputs$level2_remitted != 1) & stardOutputs$level2A_ivra_entered == 1,], table(level2A_remitted_imputed, level2A_tx, useNA = "always")))
(outputs_level2A_responded				<- with(stardOutputs[stardOutputs$meetInclusion == 1 & (is.na(stardOutputs$level1_remitted ) | stardOutputs$level1_remitted != 1) & (is.na(stardOutputs$level2_remitted ) | stardOutputs$level2_remitted != 1) & stardOutputs$level2A_ivra_entered == 1,], table(level2A_responded, level2A_tx, useNA = "always")))
(outputs_level2A_responded_imputed			<- with(stardOutputs[stardOutputs$meetInclusion == 1 & (is.na(stardOutputs$level1_remitted ) | stardOutputs$level1_remitted != 1) & (is.na(stardOutputs$level2_remitted ) | stardOutputs$level2_remitted != 1) & stardOutputs$level2A_ivra_entered == 1,], table(level2A_responded_imputed, level2A_tx, useNA = "always")))
(outputs_level2A_fu_ivra_entered			<- with(stardOutputs[stardOutputs$src_subject_id %in% level2A_entered28 & !(stardOutputs$src_subject_id %in% c(level3_entered313, 13, level4_entered94)),], table(fu_ivra_entered, level2A_tx, useNA = "always")))
(outputs_level2A_fu_relapse				<- with(stardOutputs[stardOutputs$src_subject_id %in% level2A_entered28 & !(stardOutputs$src_subject_id %in% c(level3_entered313, 13, level4_entered94)),], table(fu_relapse, level2A_tx, useNA = "always")))
(outputs_level2A_fu_susRemission_full		<- with(stardOutputs[stardOutputs$src_subject_id %in% level2A_entered28 & !(stardOutputs$src_subject_id %in% c(level3_entered313, 13, level4_entered94)),], table(fu_susRemission_full, level2A_tx, useNA = "always")))
(outputs_level2A_fu_susRemission_weak		<- with(stardOutputs[stardOutputs$src_subject_id %in% level2A_entered28 & !(stardOutputs$src_subject_id %in% c(level3_entered313, 13, level4_entered94)),], table(fu_susRemission_weak, level2A_tx, useNA = "always")))
(outputs_level2A_fu_hasAtLeast1Observations 	<- with(stardOutputs[stardOutputs$src_subject_id %in% level2A_entered28 & !(stardOutputs$src_subject_id %in% c(level3_entered313, 13, level4_entered94)),], table(fu_hasAtLeast1Observations , level2A_tx, useNA = "always")))

(outputs_level2A_fu_remissionbeforeFU			<- with(stardOutputs[stardOutputs$src_subject_id %in% level2A_entered28 & !(stardOutputs$src_subject_id %in% c(level3_entered313, 13, level4_entered94)),], table(fu_remissionbeforeFU, level2A_tx, useNA = "always")))
(outputs_level2A_fu_remissionbeforeFU_imputed		<- with(stardOutputs[stardOutputs$src_subject_id %in% level2A_entered28 & !(stardOutputs$src_subject_id %in% c(level3_entered313, 13, level4_entered94)),], table(fu_remissionbeforeFU_imputed, level2A_tx, useNA = "always")))
(outputs_level2A_fu_remitImputed_then_relapse			<- with(stardOutputs[stardOutputs$src_subject_id %in% level2A_entered28 & !(stardOutputs$src_subject_id %in% c(level3_entered313, 13, level4_entered94)),], table(fu_remitImputed_then_relapse, level2A_tx, useNA = "always")))
(outputs_level2A_fu_NOremitImputed_then_relapse 		<- with(stardOutputs[stardOutputs$src_subject_id %in% level2A_entered28 & !(stardOutputs$src_subject_id %in% c(level3_entered313, 13, level4_entered94)),], table(fu_NOremitImputed_then_relapse , level2A_tx, useNA = "always")))
(outputs_level2A_fu_remitImputed_then_susRemission_full 	<- with(stardOutputs[stardOutputs$src_subject_id %in% level2A_entered28 & !(stardOutputs$src_subject_id %in% c(level3_entered313, 13, level4_entered94)),], table(fu_remitImputed_then_susRemission_full , level2A_tx, useNA = "always")))
(outputs_level2A_fu_NOremitImputed_then_susRemission_full	<- with(stardOutputs[stardOutputs$src_subject_id %in% level2A_entered28 & !(stardOutputs$src_subject_id %in% c(level3_entered313, 13, level4_entered94)),], table(fu_NOremitImputed_then_susRemission_full, level2A_tx, useNA = "always")))
(outputs_level2A_fu_remitImputed_then_susRemission_weak	<- with(stardOutputs[stardOutputs$src_subject_id %in% level2A_entered28 & !(stardOutputs$src_subject_id %in% c(level3_entered313, 13, level4_entered94)),], table(fu_remitImputed_then_susRemission_weak, level2A_tx, useNA = "always")))
(outputs_level2A_fu_NOremitImputed_then_susRemission_weak	<- with(stardOutputs[stardOutputs$src_subject_id %in% level2A_entered28 & !(stardOutputs$src_subject_id %in% c(level3_entered313, 13, level4_entered94)),], table(fu_NOremitImputed_then_susRemission_weak, level2A_tx, useNA = "always")))

## Level 3	##
(outputs_level3_count					<- with(stardOutputs[stardOutputs$meetInclusion == 1 & (is.na(stardOutputs$level1_remitted ) | stardOutputs$level1_remitted != 1) & (is.na(stardOutputs$level2_remitted ) | stardOutputs$level2_remitted != 1) & (is.na(stardOutputs$level2A_remitted ) | stardOutputs$level2A_remitted != 1)  & stardOutputs$level3_ivra_entered == 1,], table(level3_tx, useNA = "always")))
(outputs_level3_remitted				<- with(stardOutputs[stardOutputs$meetInclusion == 1  & (is.na(stardOutputs$level1_remitted ) | stardOutputs$level1_remitted != 1) & (is.na(stardOutputs$level2_remitted ) | stardOutputs$level2_remitted != 1) & (is.na(stardOutputs$level2A_remitted ) | stardOutputs$level2A_remitted != 1)  & stardOutputs$level3_ivra_entered == 1,], table(level3_remitted, level3_tx, useNA = "always")))
(outputs_level3_remitted_imputed			<- with(stardOutputs[stardOutputs$meetInclusion == 1  & (is.na(stardOutputs$level1_remitted ) | stardOutputs$level1_remitted != 1) & (is.na(stardOutputs$level2_remitted ) | stardOutputs$level2_remitted != 1) & (is.na(stardOutputs$level2A_remitted ) | stardOutputs$level2A_remitted != 1)  & stardOutputs$level3_ivra_entered == 1,], table(level3_remitted_imputed, level3_tx, useNA = "always")))
(outputs_level3_responded				<- with(stardOutputs[stardOutputs$meetInclusion == 1  & (is.na(stardOutputs$level1_remitted ) | stardOutputs$level1_remitted != 1) & (is.na(stardOutputs$level2_remitted ) | stardOutputs$level2_remitted != 1) & (is.na(stardOutputs$level2A_remitted ) | stardOutputs$level2A_remitted != 1)  & stardOutputs$level3_ivra_entered == 1,], table(level3_responded, level3_tx, useNA = "always")))
(outputs_level3_responded_imputed			<- with(stardOutputs[stardOutputs$meetInclusion == 1  & (is.na(stardOutputs$level1_remitted ) | stardOutputs$level1_remitted != 1) & (is.na(stardOutputs$level2_remitted ) | stardOutputs$level2_remitted != 1) & (is.na(stardOutputs$level2A_remitted ) | stardOutputs$level2A_remitted != 1)  & stardOutputs$level3_ivra_entered == 1,], table(level3_responded_imputed, level3_tx, useNA = "always")))
(outputs_level3_fu_ivra_entered			<- with(stardOutputs[stardOutputs$src_subject_id %in% level3_entered313 & !(stardOutputs$src_subject_id %in% c(level4_entered94)),], table(fu_ivra_entered, level3_tx, useNA = "always")))
(outputs_level3_fu_relapse				<- with(stardOutputs[stardOutputs$src_subject_id %in% level3_entered313 & !(stardOutputs$src_subject_id %in% c(level4_entered94)),], table(fu_relapse, level3_tx, useNA = "always")))
(outputs_level3_fu_susRemission_full		<- with(stardOutputs[stardOutputs$src_subject_id %in% level3_entered313 & !(stardOutputs$src_subject_id %in% c(level4_entered94)),], table(fu_susRemission_full, level3_tx, useNA = "always")))
(outputs_level3_fu_susRemission_weak		<- with(stardOutputs[stardOutputs$src_subject_id %in% level3_entered313 & !(stardOutputs$src_subject_id %in% c(level4_entered94)),], table(fu_susRemission_weak, level3_tx, useNA = "always")))
(outputs_level3_fu_hasAtLeast1Observations 	<- with(stardOutputs[stardOutputs$src_subject_id %in% level3_entered313 & !(stardOutputs$src_subject_id %in% c(level4_entered94)),], table(fu_hasAtLeast1Observations , level3_tx, useNA = "always")))

(outputs_level3_fu_remissionbeforeFU			<- with(stardOutputs[stardOutputs$src_subject_id %in% level3_entered313 & !(stardOutputs$src_subject_id %in% c(level4_entered94)),], table(fu_remissionbeforeFU, level3_tx, useNA = "always")))
(outputs_level3_fu_remissionbeforeFU_imputed		<- with(stardOutputs[stardOutputs$src_subject_id %in% level3_entered313 & !(stardOutputs$src_subject_id %in% c(level4_entered94)),], table(fu_remissionbeforeFU_imputed, level3_tx, useNA = "always")))
(outputs_level3_fu_remitImputed_then_relapse			<- with(stardOutputs[stardOutputs$src_subject_id %in% level3_entered313 & !(stardOutputs$src_subject_id %in% c(level4_entered94)),], table(fu_remitImputed_then_relapse, level3_tx, useNA = "always")))
(outputs_level3_fu_NOremitImputed_then_relapse 		<- with(stardOutputs[stardOutputs$src_subject_id %in% level3_entered313 & !(stardOutputs$src_subject_id %in% c(level4_entered94)),], table(fu_NOremitImputed_then_relapse , level3_tx, useNA = "always")))
(outputs_level3_fu_remitImputed_then_susRemission_full 	<- with(stardOutputs[stardOutputs$src_subject_id %in% level3_entered313 & !(stardOutputs$src_subject_id %in% c(level4_entered94)),], table(fu_remitImputed_then_susRemission_full , level3_tx, useNA = "always")))
(outputs_level3_fu_NOremitImputed_then_susRemission_full	<- with(stardOutputs[stardOutputs$src_subject_id %in% level3_entered313 & !(stardOutputs$src_subject_id %in% c(level4_entered94)),], table(fu_NOremitImputed_then_susRemission_full, level3_tx, useNA = "always")))
(outputs_level3_fu_remitImputed_then_susRemission_weak	<- with(stardOutputs[stardOutputs$src_subject_id %in% level3_entered313 & !(stardOutputs$src_subject_id %in% c(level4_entered94)),], table(fu_remitImputed_then_susRemission_weak, level3_tx, useNA = "always")))
(outputs_level3_fu_NOremitImputed_then_susRemission_weak	<- with(stardOutputs[stardOutputs$src_subject_id %in% level3_entered313 & !(stardOutputs$src_subject_id %in% c(level4_entered94)),], table(fu_NOremitImputed_then_susRemission_weak, level3_tx, useNA = "always")))

## Level 4	##
(outputs_level4_count					<- with(stardOutputs[stardOutputs$meetInclusion == 1 & (is.na(stardOutputs$level1_remitted ) | stardOutputs$level1_remitted != 1) & (is.na(stardOutputs$level2_remitted ) | stardOutputs$level2_remitted != 1) & (is.na(stardOutputs$level2A_remitted ) | stardOutputs$level2A_remitted != 1)  & (is.na(stardOutputs$level3_remitted ) | stardOutputs$level3_remitted != 1)  & stardOutputs$level4_ivra_entered == 1,], table(level4_tx, useNA = "always")))
(outputs_level4_remitted				<- with(stardOutputs[stardOutputs$meetInclusion == 1 & (is.na(stardOutputs$level1_remitted ) | stardOutputs$level1_remitted != 1) & (is.na(stardOutputs$level2_remitted ) | stardOutputs$level2_remitted != 1) & (is.na(stardOutputs$level2A_remitted ) | stardOutputs$level2A_remitted != 1)  & (is.na(stardOutputs$level3_remitted ) | stardOutputs$level3_remitted != 1)  & stardOutputs$level4_ivra_entered == 1,], table(level4_remitted, level4_tx, useNA = "always")))
(outputs_level4_remitted_imputed			<- with(stardOutputs[stardOutputs$meetInclusion == 1 & (is.na(stardOutputs$level1_remitted ) | stardOutputs$level1_remitted != 1) & (is.na(stardOutputs$level2_remitted ) | stardOutputs$level2_remitted != 1) & (is.na(stardOutputs$level2A_remitted ) | stardOutputs$level2A_remitted != 1)  & (is.na(stardOutputs$level3_remitted ) | stardOutputs$level3_remitted != 1)  & stardOutputs$level4_ivra_entered == 1,], table(level4_remitted_imputed, level4_tx, useNA = "always")))
(outputs_level4_responded				<- with(stardOutputs[stardOutputs$meetInclusion == 1 & (is.na(stardOutputs$level1_remitted ) | stardOutputs$level1_remitted != 1) & (is.na(stardOutputs$level2_remitted ) | stardOutputs$level2_remitted != 1) & (is.na(stardOutputs$level2A_remitted ) | stardOutputs$level2A_remitted != 1)  & (is.na(stardOutputs$level3_remitted ) | stardOutputs$level3_remitted != 1)  & stardOutputs$level4_ivra_entered == 1,], table(level4_responded, level4_tx, useNA = "always")))
(outputs_level4_responded_imputed			<- with(stardOutputs[stardOutputs$meetInclusion == 1 & (is.na(stardOutputs$level1_remitted ) | stardOutputs$level1_remitted != 1) & (is.na(stardOutputs$level2_remitted ) | stardOutputs$level2_remitted != 1) & (is.na(stardOutputs$level2A_remitted ) | stardOutputs$level2A_remitted != 1)  & (is.na(stardOutputs$level3_remitted ) | stardOutputs$level3_remitted != 1)  & stardOutputs$level4_ivra_entered == 1,], table(level4_responded_imputed, level4_tx, useNA = "always")))
(outputs_level4_fu_ivra_entered			<- with(stardOutputs[stardOutputs$src_subject_id %in% level4_entered94,], table(fu_ivra_entered, level4_tx, useNA = "always")))
(outputs_level4_fu_relapse				<- with(stardOutputs[stardOutputs$src_subject_id %in% level4_entered94,], table(fu_relapse, level4_tx, useNA = "always")))
(outputs_level4_fu_susRemission_full		<- with(stardOutputs[stardOutputs$src_subject_id %in% level4_entered94,], table(fu_susRemission_full, level4_tx, useNA = "always")))
(outputs_level4_fu_susRemission_weak		<- with(stardOutputs[stardOutputs$src_subject_id %in% level4_entered94,], table(fu_susRemission_weak, level4_tx, useNA = "always")))
(outputs_level4_fu_hasAtLeast1Observations 		<- with(stardOutputs[stardOutputs$src_subject_id %in% level4_entered94,], table(fu_hasAtLeast1Observations , level4_tx, useNA = "always")))

(outputs_level4_fu_remissionbeforeFU			<- with(stardOutputs[stardOutputs$src_subject_id %in% level4_entered94,], table(fu_remissionbeforeFU, level4_tx, useNA = "always")))
(outputs_level4_fu_remissionbeforeFU_imputed		<- with(stardOutputs[stardOutputs$src_subject_id %in% level4_entered94,], table(fu_remissionbeforeFU_imputed, level4_tx, useNA = "always")))

(outputs_level4_fu_remitImputed_then_relapse			<- with(stardOutputs[stardOutputs$src_subject_id %in% level4_entered94,], table(fu_remitImputed_then_relapse, level4_tx, useNA = "always")))
(outputs_level4_fu_NOremitImputed_then_relapse 		<- with(stardOutputs[stardOutputs$src_subject_id %in% level4_entered94,], table(fu_NOremitImputed_then_relapse , level4_tx, useNA = "always")))
(outputs_level4_fu_remitImputed_then_susRemission_full 	<- with(stardOutputs[stardOutputs$src_subject_id %in% level4_entered94,], table(fu_remitImputed_then_susRemission_full , level4_tx, useNA = "always")))
(outputs_level4_fu_NOremitImputed_then_susRemission_full	<- with(stardOutputs[stardOutputs$src_subject_id %in% level4_entered94,], table(fu_NOremitImputed_then_susRemission_full, level4_tx, useNA = "always")))
(outputs_level4_fu_remitImputed_then_susRemission_weak	<- with(stardOutputs[stardOutputs$src_subject_id %in% level4_entered94,], table(fu_remitImputed_then_susRemission_weak, level4_tx, useNA = "always")))
(outputs_level4_fu_NOremitImputed_then_susRemission_weak	<- with(stardOutputs[stardOutputs$src_subject_id %in% level4_entered94,], table(fu_NOremitImputed_then_susRemission_weak, level4_tx, useNA = "always")))

## Function to extract attributes from output table	##
extract <- function(outputTable){
	rownames(outputTable) <- paste(names(attributes(outputTable)$dimnames)[[1]], rownames(outputTable), sep = " ")
	return(outputTable)
}

### Merging and savings outputs 	###

summaryLevel1 <- rbind(outputs_level1_count, extract( outputs_level1_remitted), extract(outputs_level1_remitted_imputed), extract( outputs_level1_responded), extract(outputs_level1_responded_imputed), extract(outputs_level1_fu_ivra_entered),  extract( outputs_level1_fu_remissionbeforeFU), extract( outputs_level1_fu_remissionbeforeFU_imputed),extract( outputs_level1_fu_relapse), extract( outputs_level1_fu_susRemission_full), extract(outputs_level1_fu_susRemission_weak), extract(outputs_level1_fu_hasAtLeast1Observations), extract(outputs_level1_fu_remitImputed_then_relapse),extract(outputs_level1_fu_NOremitImputed_then_relapse), extract(outputs_level1_fu_remitImputed_then_susRemission_full), extract(outputs_level1_fu_NOremitImputed_then_susRemission_full),	extract(outputs_level1_fu_remitImputed_then_susRemission_weak),	extract(outputs_level1_fu_NOremitImputed_then_susRemission_weak))
summaryLevel2 <- rbind(outputs_level2_count, extract( outputs_level2_remitted), extract(outputs_level2_remitted_imputed), extract( outputs_level2_responded), extract(outputs_level2_responded_imputed), extract(outputs_level2_fu_ivra_entered),  extract( outputs_level2_fu_remissionbeforeFU), extract( outputs_level2_fu_remissionbeforeFU_imputed),extract( outputs_level2_fu_relapse), extract( outputs_level2_fu_susRemission_full), extract(outputs_level2_fu_susRemission_weak), extract(outputs_level2_fu_hasAtLeast1Observations), extract(outputs_level2_fu_remitImputed_then_relapse),extract(outputs_level2_fu_NOremitImputed_then_relapse), extract(outputs_level2_fu_remitImputed_then_susRemission_full), extract(outputs_level2_fu_NOremitImputed_then_susRemission_full),	extract(outputs_level2_fu_remitImputed_then_susRemission_weak),	extract(outputs_level2_fu_NOremitImputed_then_susRemission_weak))
summaryLevel2A <- rbind(outputs_level2A_count, extract( outputs_level2A_remitted), extract(outputs_level2A_remitted_imputed), extract( outputs_level2A_responded), extract(outputs_level2A_responded_imputed), extract(outputs_level2A_fu_ivra_entered),  extract( outputs_level2A_fu_remissionbeforeFU), extract( outputs_level2A_fu_remissionbeforeFU_imputed),extract( outputs_level2A_fu_relapse), extract( outputs_level2A_fu_susRemission_full), extract(outputs_level2A_fu_susRemission_weak), extract(outputs_level2A_fu_hasAtLeast1Observations), extract(outputs_level2A_fu_remitImputed_then_relapse),extract(outputs_level2A_fu_NOremitImputed_then_relapse), extract(outputs_level2A_fu_remitImputed_then_susRemission_full), extract(outputs_level2A_fu_NOremitImputed_then_susRemission_full),	extract(outputs_level2A_fu_remitImputed_then_susRemission_weak),	extract(outputs_level2A_fu_NOremitImputed_then_susRemission_weak))
summaryLevel3 <- rbind(outputs_level3_count, extract( outputs_level3_remitted), extract(outputs_level3_remitted_imputed), extract( outputs_level3_responded), extract(outputs_level3_responded_imputed), extract(outputs_level3_fu_ivra_entered),  extract( outputs_level3_fu_remissionbeforeFU), extract( outputs_level3_fu_remissionbeforeFU_imputed),extract( outputs_level3_fu_relapse), extract( outputs_level3_fu_susRemission_full), extract(outputs_level3_fu_susRemission_weak), extract(outputs_level3_fu_hasAtLeast1Observations), extract(outputs_level3_fu_remitImputed_then_relapse),extract(outputs_level3_fu_NOremitImputed_then_relapse), extract(outputs_level3_fu_remitImputed_then_susRemission_full), extract(outputs_level3_fu_NOremitImputed_then_susRemission_full),	extract(outputs_level3_fu_remitImputed_then_susRemission_weak),	extract(outputs_level3_fu_NOremitImputed_then_susRemission_weak))
summaryLevel4 <- rbind(outputs_level4_count, extract( outputs_level4_remitted), extract(outputs_level4_remitted_imputed), extract( outputs_level4_responded), extract(outputs_level4_responded_imputed), extract(outputs_level4_fu_ivra_entered),  extract( outputs_level4_fu_remissionbeforeFU), extract( outputs_level4_fu_remissionbeforeFU_imputed),extract( outputs_level4_fu_relapse), extract( outputs_level4_fu_susRemission_full), extract(outputs_level4_fu_susRemission_weak), extract(outputs_level4_fu_hasAtLeast1Observations), extract(outputs_level4_fu_remitImputed_then_relapse),extract(outputs_level4_fu_NOremitImputed_then_relapse), extract(outputs_level4_fu_remitImputed_then_susRemission_full), extract(outputs_level4_fu_NOremitImputed_then_susRemission_full),	extract(outputs_level4_fu_remitImputed_then_susRemission_weak),	extract(outputs_level4_fu_NOremitImputed_then_susRemission_weak))

### Outputting HRSD Change

hrsdChangeLevel1 <- with(stardOutputs[stardOutputs$meetInclusion == 1 & stardOutputs$level1_ivra_entered == 1,], aggregate(level1_hrsdchange_imputed, list(level1_tx), summarizeOutputs))
hrsdChangeLevel2 <- with(stardOutputs[stardOutputs$meetInclusion == 1 & (is.na(stardOutputs$level1_remitted ) | stardOutputs$level1_remitted != 1) & stardOutputs$level2_ivra_entered == 1,], aggregate(level2_hrsdchange_imputed, list(level2_tx),  summarizeOutputs))
hrsdChangeLevel2A <- with(stardOutputs[stardOutputs$meetInclusion == 1 & (is.na(stardOutputs$level1_remitted ) | stardOutputs$level1_remitted != 1) & (is.na(stardOutputs$level2_remitted ) | stardOutputs$level2_remitted != 1) & stardOutputs$level2A_ivra_entered == 1,], aggregate(level2A_hrsdchange_imputed, list(level2A_tx),  summarizeOutputs))
hrsdChangeLevel3 <- with(stardOutputs[stardOutputs$meetInclusion == 1  & (is.na(stardOutputs$level1_remitted ) | stardOutputs$level1_remitted != 1) & (is.na(stardOutputs$level2_remitted ) | stardOutputs$level2_remitted != 1) & (is.na(stardOutputs$level2A_remitted ) | stardOutputs$level2A_remitted != 1)  & stardOutputs$level3_ivra_entered == 1,], aggregate(level3_hrsdchange_imputed, list(level3_tx),  summarizeOutputs))
hrsdChangeLevel4 <- with(stardOutputs[stardOutputs$meetInclusion == 1 & (is.na(stardOutputs$level1_remitted ) | stardOutputs$level1_remitted != 1) & (is.na(stardOutputs$level2_remitted ) | stardOutputs$level2_remitted != 1) & (is.na(stardOutputs$level2A_remitted ) | stardOutputs$level2A_remitted != 1)  & (is.na(stardOutputs$level3_remitted ) | stardOutputs$level3_remitted != 1)  & stardOutputs$level4_ivra_entered == 1,], aggregate(level4_hrsdchange_imputed, list(level4_tx),  summarizeOutputs))

write.csv(t(summaryLevel1), "OutputFolder/STARD Summary Level1.csv")
write.csv(t(summaryLevel2), "OutputFolder/STARD Summary Level2.csv")
write.csv(t(summaryLevel2A), "OutputFolder/STARD Summary Level2A.csv")
write.csv(t(summaryLevel3), "OutputFolder/STARD Summary Level3.csv")
write.csv(t(summaryLevel4), "OutputFolder/STARD Summary Level4.csv")

write.csv(t(hrsdChangeLevel1 ), "OutputFolder/STARD hrsdChange Level1.csv")
write.csv(t(hrsdChangeLevel2 ), "OutputFolder/STARD hrsdChange Level2.csv")
write.csv(t(hrsdChangeLevel2A ), "OutputFolder/STARD hrsdChange Level2A.csv")
write.csv(t(hrsdChangeLevel3 ), "OutputFolder/STARD hrsdChange Level3.csv")
write.csv(t(hrsdChangeLevel4 ), "OutputFolder/STARD hrsdChange Level4.csv")

write.csv(stardOutputs, "OutputFolder/STARD - patientOutputs.csv")
```

**Supplement 7:**

**Demographics and clinical characteristics**

|  | **Bupropion**  **(n=190)** | | **Sertraline**  **(n=198)** | | **Venlafaxine**  **(n=192)** | | **Total Switch (n=580)** | |
| --- | --- | --- | --- | --- | --- | --- | --- | --- |
| **Demographic** | **Mean** | **SD** | **Mean** | **SD** | **Mean** | **SD** | **Mean** | **SD** |
| Age | 43.0 | 13.2 | 42.4 | 12.4 | 41.6 | 12.7 | 42.3 | 12.7 |
| Education (years) | 13.4 | 2.8 | 13.1 | 3.2 | 13.3 | 3.0 | 13.3 | 3.0 |
| Monthly household income | 2000.9 | 2645.6 | 1888.6 | 2731.8 | 1892.9 | 1904.5 | 1926.1 | 2456.0 |
|  | **n** | **%** | **n** | **%** | **n** | **%** | **n** | **%** |
| Female | 108 | 56.8 | 111 | 56.1 | 122 | 63.5 | 341 | 58.8 |
| Race |  |  |  |  |  |  |  |  |
| White | 138 | 72.6 | 155 | 78.3 | 138 | 71.9 | 431 | 74.3 |
| Black | 24 | 12.6 | 20 | 10.1 | 25 | 13.0 | 69 | 11.9 |
| Other | 28 | 14.7 | 23 | 11.6 | 29 | 15.1 | 80 | 13.8 |
| Hispanic or Latino | 17 | 8.9 | 26 | 13.1 | 23 | 12.0 | 66 | 11.4 |
| Employment status |  |  |  |  |  |  |  |  |
| Employed | 99 | 52.1 | 97 | 49.0 | 99 | 51.6 | 295 | 50.9 |
| Unemployed | 75 | 39.5 | 92 | 46.5 | 81 | 42.2 | 248 | 42.8 |
| Retired | 15 | 7.9 | 8 | 4.0 | 9 | 4.7 | 32 | 5.5 |
| Medical insurance |  |  |  |  |  |  |  |  |
| Private | 80 | 42.1 | 80 | 40.4 | 84 | 43.8 | 244 | 42.1 |
| Public | 46 | 24.2 | 32 | 16.2 | 35 | 18.2 | 113 | 19.5 |
| None | 70 | 36.8 | 88 | 44.4 | 75 | 39.1 | 233 | 40.2 |
| Marital status |  |  |  |  |  |  |  |  |
| Single | 56 | 29.5 | 57 | 28.8 | 53 | 27.6 | 166 | 28.6 |
| Married/cohabiting | 73 | 38.4 | 86 | 43.4 | 77 | 40.1 | 236 | 40.7 |
| Divorce/separated | 51 | 26.8 | 46 | 23.2 | 55 | 28.6 | 152 | 26.2 |
| Widowed | 10 | 5.3 | 9 | 4.5 | 7 | 3.6 | 26 | 4.5 |
| **Clinical Features** | **n** | **%** | **n** | **%** | **n** | **%** | **n** | **%** |
| First episode occurrence before age 18 | 74 | 39.2 | 74 | 38.1 | 70 | 37.0 | 218 | 38.1 |
| Recurrent MDD | 110 | 64.3 | 118 | 65.6 | 118 | 67.4 | 346 | 65.8 |
| Family history of depression | 102 | 54.3 | 105 | 54.7 | 98 | 53.0 | 305 | 54.0 |
| Prior suicide attempt | 34 | 18.1 | 40 | 20.2 | 33 | 17.2 | 107 | 18.5 |
| Duration of current episode ≥ 2 years | 54 | 28.6 | 55 | 28.4 | 53 | 28.0 | 162 | 28.3 |
|  | **Mean** | **SD** | **Mean** | **SD** | **Mean** | **SD** | **Mean** | **SD** |
| Age at first episode (years) | 25.8 | 15.0 | 24.2 | 13.2 | 24.5 | 13.7 | 24.8 | 14.0 |
| Illness duration (years) | 17.3 | 14.2 | 18.5 | 14.4 | 17.1 | 13.6 | 17.7 | 14.1 |
| Number of MDD episodes | 7.1 | 12.2 | 6.0 | 10.6 | 7.8 | 12.8 | 7.0 | 11.9 |
| Duration of current episode (months) | 36.0 | 81.9 | 27.1 | 57.0 | 26.9 | 53.1 | 30.0 | 65.2 |
| Median duration of current episode (months) | 8.9 |  | 10.7 |  | 8.9 |  | 9.5 |  |
| HRSD_17_ score (at entry into step-2) | 20.5 | 6.4 | 20.2 | 6.0 | 20.2 | 6.1 | 20.3 | 6.2 |
| Cumulative Illness Rating Scale |  |  |  |  |  |  |  |  |
| Categories endorsed | 2.7 | 1.6 | 2.7 | 1.7 | 2.8 | 1.5 | 2.8 | 1.6 |
| Total score | 5.4 | 4.3 | 5.2 | 4.0 | 5.5 | 4.0 | 5.3 | 4.1 |
| Severity score | 1.8 | 0.7 | 1.9 | 1.0 | 1.8 | 0.7 | 1.9 | 0.8 |

* Note that sums do not always equal n due to missing values. Percentages are based on available data.

**Supplement 8:**

**Primary Outcome Remission Rate Comparison**

| Step-2  Drug-Switch Treatments | Primary Outcome as Reported in NEJM:  % HRSD Remissions | Primary Outcome as Reported in RIAT Reanalysis:  % HRSD Remissions |
| --- | --- | --- |
| BUP | 21.3% | 16.3% |
| SER | 17.6% | 16.2% |
| VEN | 24.8% | 19.3% |
| **Average Switch Treatment** | **21.2%** | **17.5%** |

**Supplement 9:**

**TESI Rates for Step-2 Drug-Switch and Step-1 Citalopram Treatments**

|  | **BUP** | **SER** | **VEN** | **Total**  **Drug-Switch** | **Step-1 Citalopram** |
| --- | --- | --- | --- | --- | --- |
| **Number of evaluable patients** | 190 | 198 | 192 | 580 | 3,110 |
| **Number of evaluable patients with a baseline QIDS-C suicide ideation score of 0 or 1. These patients are classified as Non-Suicidal at baseline.** | 134 | 143 | 133 | 410 | 2,123 |
| **Number of Non-Suicidal patients at baseline with one or more post-baseline QIDS-C suicide ideation scores of 2 or 3—This is the number of TESI patients.** | 17 | 16 | 20 | 53 | 190 |
| **TESI Rate (Divide the number of TESI patients by number of patients classified as Non-Suicidal at baseline)** | 12.7% | 11.2% | 15.0% | 12.9% | 9.0% |
